# Power Laws in Superspreading Events: Evidence from Coronavirus Outbreaks and Implications for SIR Models

**DOI:** 10.1101/2020.06.11.20128058

**Authors:** Masao Fukui, Chishio Furukawa

## Abstract

While they are rare, superspreading events (SSEs), wherein a few primary cases infect an extraordinarily large number of secondary cases, are recognized as a prominent determinant of aggregate infection rates (ℛ_0_). Existing stochastic SIR models incorporate SSEs by fitting distributions with thin tails, or finite variance, and therefore predicting almost deterministic epidemiological outcomes in large populations. This paper documents evidence from recent coronavirus outbreaks, including SARS, MERS, and COVID-19, that SSEs follow a power law distribution with fat tails, or infinite variance. We then extend an otherwise standard SIR model with the estimated power law distributions, and show that idiosyncratic uncertainties in SSEs will lead to large aggregate uncertainties in infection dynamics, even with large populations. That is, the timing and magnitude of outbreaks will be unpredictable. While such uncertainties have social costs, we also find that they on average *decrease* the herd immunity thresholds and the cumulative infections because per-period infection rates have decreasing marginal effects. Our findings have implications for social distancing interventions: targeting SSEs reduces not only the average rate of infection (ℛ_0_) but also its uncertainty. To understand this effect, and to improve inference of the average reproduction numbers under fat tails, estimating the tail distribution of SSEs is vital.

## 1 Introduction

On March 10th, 2020, choir members were gathered for their rehearsal in Washington. While they were all cautious to keep distance from one another and nobody was coughing, three weeks later, 52 members had COVID-19, and two passed away. There are numerous similar anecdotes worldwide.^1^ Many studies have shown that the average basic reproduction number (ℛ_0_) is around 2.5-3 for this coronavirus (e.g. Liu et al., 2020), but 75% of infected cases do not pass on to any others (Nishiura et al., 2020). The superspreading events (SSEs), wherein a few primary cases infect an extraordinarily large number of others, are responsible for the high average number. As SSEs were also prominent in SARS and MERS before COVID-19, epidemiology research has long sought to understand them (e.g. Shen et al., 2004). In particular, various parametric distributions of infection rates have been proposed, and their variances have been estimated in many epidemics under an assumption that they exist (e.g. Lloyd-Smith et al., 2005). On the other hand, stochastic Susceptible-Infectious-Recovered (SIR) models have shown that, as long as the infected population is moderately large, the idiosyncratic uncertainties of SSEs will cancel out each other. That is, following the Central Limit Theorem (CLT), stochastic models quickly converge to their deterministic counterparts, and become largely predictable. From this perspective, the dispersion of SSEs is unimportant in itself, but is useful only to the extent it can help target lockdown policies to focus on SSEs to efficiently reduce the average rates ℛ_0_ (Endo et al., 2020).

In this paper, we extend this research by closely examining the distribution of infection rates, and rethinking how its dispersion influences the uncertainties of aggregate dynamics. Using evidence from the several coronavirus outbreaks, we show that SSEs follow a power law, or Pareto, distribution with fat tails, or infinite variance. That is, the true variance of infection rates cannot be empirically estimated as any estimate will be an underestimate however large it may be. When the CLT assumption of finite variance does not hold, many theoretical and statistical implications of epidemiology models will require rethinking. Theoretically, even when the infected population is large, the idiosyncratic uncertainties in SSEs will persist and lead to large aggregate uncertainties. Statistically, the standard estimate of the average reproduction number (ℛ_0_) may be far from its true mean, and the standard errors will understate the true uncertainty. Because the infected population for COVID-19 is already large, our findings have immediate implications for statistical inference and current policy.

We begin with evidence. Figure 1 plots the largest clusters reported worldwide for COVID-19 from data gathered by Leclerc et al. (2020). If a random variable follows a power law distribution with an exponent *α*, then the log of its scale (e.g. a US navy vessel had 1,156 cases tested positive) and the log of its severity rank (e.g. that navy case ranked 1st in severity) will have a linear relationship, with its slope indicating −*α*. Figure 1 shows a fine fit of the power law distribution (*R*^2^ = 0.98).^2^ Moreover, the slope is very close to 1, indicating a significant fatness of the tail to the extent that is analogous to natural disasters such as earthquakes (Gutenberg and Richter, 1954) that are infrequent but can be extreme. While data collection through media reports may be biased towards extreme cases, analogous relationships hold for SARS, MERS, and COVID-19 data based on surveillance data, with exponents often indicating fat tails. Note that other distributions, including the negative binomial distributions commonly applied in epidemiology research, cannot predict these relationships, and significantly underestimate the risks of extremely severe SSEs.

**Figure 1:**
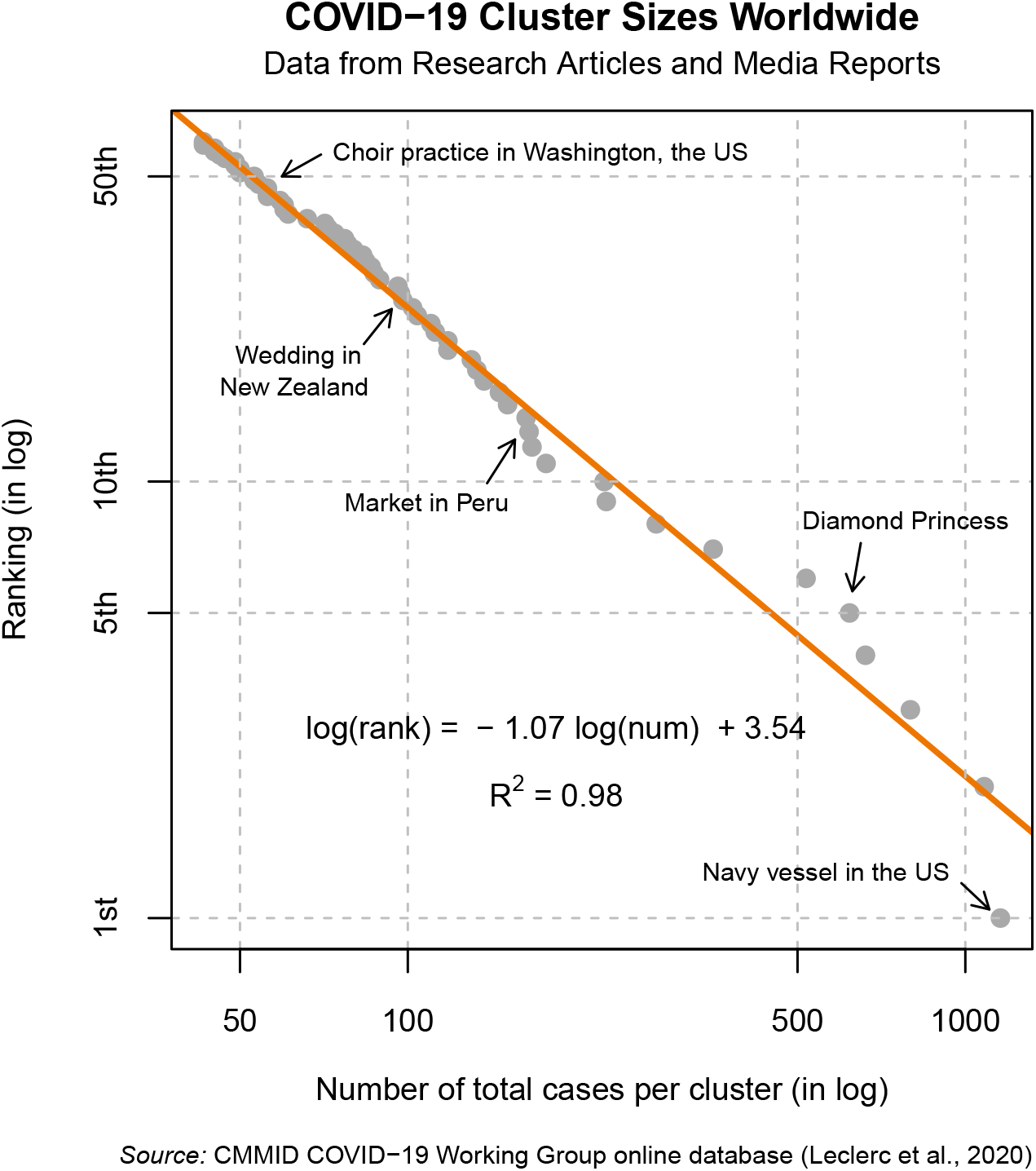
Log cluster size vs log rank for COVID-19 worldwide. *Notes*: Figure 1 plots the number of total cases per cluster (in log) and their ranks (in log) for COVID-19, last updated on June 3rd. It fits a linear regression for the clusters with size larger than 40. The data are collected by the Centre for the Mathematical Modelling of Infectious Diseases COVID-19 Working Group (Leclerc et al., 2020).

Using estimated power law distributions, we show that stochastic SIR models predict substantial uncertainties in aggregate epidemiological outcomes. Concretely, we consider a stochastic model with a population of one million, whereby a thousand people are initially infected, and apply epidemiological parameters adopted from the literature. We consider effects of tails of distribution while keeping the average rate (ℛ_0_) constant. Under thin-tailed distributions, such as the estimated negative binomial distribution or power law distribution with *α* = 2, the epidemiological outcomes will be essentially predictable. However, under fat-tailed distributions as estimated in the COVID-19 data worldwide (*α* = 1.1), there will be immense variations in all outcomes. For example, the peak infection rate is on average 19%, but its 90the percentile is 34% while its 10th percentile is 10%. Under thin-tailed distribution such as negative binomial distribution, the average, 90th percentile and 10th percentile of the peak infection is all concentrated at 27%, generating almost deterministic outcomes.

While our primary focus was on the effect on aggregate uncertainty, we also find important effects on average outcomes. In particular, under a fat-tailed distribution, the cumulative and peak infection, as well as the herd immunity threshold, will be lower, and the timing of outbreak will come later than those under a thin-tailed distribution, *on average*. For example, the average herd immunity threshold is 65 percent with thin-tailed distribution, it is 47% with fat-tailed distribution. These observations suggest that the increase in aggregate uncertainty over ℛ_0_ has effects analogous to a decrease in average ℛ_0_. This relationship arises because the average future infection will be a *concave* function of today’s infection rate: because of concavity, mean preserving spread will lower the average level. In particular, today’s higher infection rate has two countering effects: while it increases the future infection, it also decreases the susceptible population, which decreases it. We provide theoretical interpretations for each outcome by examining the effect of mean-preserving spread of ℛ_0_ in analytical results derived in deterministic models.

Our findings have critical implications for the design of lockdown policies to minimize the social costs of infection. Here, we study lockdown policies that target SSEs. We assume that the maximum size of infection rate can be limited to the top 5 percent with some probabilities by banning large gatherings. Because both the uncertainty and mean of the infection rate in the fat-tailed distribution are driven by the tail events, such policies substantially lower the uncertainty and improve the average outcomes. Because the cost of such policy^3^ is difficult to estimate reliably, we do not compute the cost-effectiveness of such policy. Nonetheless, we believe this is an important consideration in the current debates on how to re-open the economy while mitigating the uncertainties of subsequent waves.

Finally, we also show the implications of a fat-tailed distributions for the estimation of the average infection rate. Under such a distribution with small sample sizes, the sample mean yields estimates that are far from the true mean and standard errors that are too small. To address such possibility, it will be helpful to estimate the power law exponent. If the estimate indicates a thin-tailed distribution, then one can be confident with the sample mean estimate. If it indicates a fat-tailed distribution, then one must be aware that there is much uncertainty in the estimate not captured by its confidence interval. While such fat-tailed distributions cause notoriously difficult estimation problems, we explore a “plug-in” method that uses the estimated exponent. Such estimators generate median estimates closer to the true mean with adequate confidence intervals that reflect the substantial risk of SSEs.

## Related Literature

First, our paper belongs to a large literature on stochastic epidemiological models. The deterministic SIR model was initiated by Kermack and McKendrick (1927), and later, Bartlett (1949) and Kendall (1956) developed stochastic SIR models (see Britton (2010, 2018) for surveys). The traditional view of the stochastic SIR model is that while useful when the number of infected is small, once the infected population is moderately large, it behaves similarly to the deterministic model due to the CLT. Britton (2010) writes “Once a large number of individuals have been infected, the epidemic process may be approximated by the deterministic counter-part.” There are recent applications of stochastic SIR models that study the very beginning of COVID-19 outbreaks (for example, Abbott et al. (2020), Karako et al. (2020), Simha et al. (2020) and Bardina et al. (2020)). However, the major modeling effort has been to use deterministic models based on the common justification above. Our point is that when the distribution is fat-tailed, which we found an empirical support for, the CLT no longer applies, and hence the stochastic model behaves qualitatively differently from its deterministic counterpart even with a large number of infected individuals.

Second, the empirical importance of SSEs is widely recognized in the epidemiological literature before COVID-19 (Lloyd-Smith et al., 2005; Galvani and May, 2005) and for COVID-19 (Frieden and Lee, 2020; Endo et al., 2020). These papers fit the parametric distribution that is by construction thin-tailed, such as negative binomial distribution. It has been common to estimate “the dispersion parameter *k*” of the negative binomial distribution. We argue that the fat-tailed distribution provides a better fit to the empirical distribution of SSEs, in which a tail parameter, *α*, parsimoniously captures the thickness of the tail. A recent contribution by Cooper et al. (2019) consider Pareto rule in the context of malaria transmission, but they nonetheless estimate the dispersion with finite variance for the entire infections.

Third, our paper also relates to studies that incorporate heterogeneity into SIR models, incorporating differences in individual characteristics or community structures. Several recent papers point out that the permanent heterogeneity in individual infection rates lower the herd immunity threshold (Gomes et al., 2020; Hébert-Dufresne et al., 2020; Britton et al., 2020). Although we obtain a similar result, our underlying mechanisms are distinct from theirs. In our model, there is no ex-ante heterogeneity across individuals, and thus their mechanism is not present. Instead, what matters for us is the aggregate fluctuations in ℛ_0_, which their models do not exhibit. Some recent papers emphasize the importance of age-dependent heterogeneity and its implications for lockdown policies (Acemoglu et al., 2020; Davies et al., 2020; Gollier, 2020; Rampini, 2020; Glover et al., 2020; Brotherhood et al., 2020). We emphasize another dimension of targeting: targeting toward large social gatherings, and this policy reduces the uncertainty regarding various epidemiological outcomes. Another related paper is Beare and Toda (2020). They document that the cumulative number of infected population across cities and countries is closely approximated by a power law distribution. They then argue that the standard SIR model is able to explain the fact. We document that the infection at the individual level follows a power law.

Finally, it is well-known that many variables follow a power law distribution. These include the city size (Zipf, 1949), the firm size (Axtell, 2001), income (Atkinson et al., 2011), wealth (Kleiber and Kotz, 2003), consumption (Toda and Walsh, 2015) and even the size of the earthquakes (Gutenberg and Richter, 1954), the moon craters and solar flares (Newman, 2005). Regarding COVID-19, Beare and Toda (2020) document that the cumulative number of infected population across cities and countries is closely approximated by a power law distribution. They then argue that the standard SIR model is able to explain the fact. We document that the infection at the individual level follows a power law. We are also partly inspired by economics literature which argue that the fat-tailed distribution in firm-size has an important consequence for the macroeconomics dynamics, originated by Gabaix (2011). We follow the similar route in documenting that the SSEs are well approximated by a power law distribution and arguing that such empirical regularities have important consequences for the epidemiological dynamics.

### Roadmap

The rest of the paper is organized as follows. Section 2 documents evidence that the distribution of SSEs follows power law. Section 3 embed the evidence into an otherwise standard SIR models to demonstrate its implications for the epidemiological dynamics. Section 4 studies estimation of the reproduction numbers under fat-tailed distribution. Section 5 concludes by discussing what our results imply for ongoing COVID-19 pandemic.

## 2 Evidence

We present evidence from SARS, MERS, and COVID-19 that the SSEs follow power law distributions. Moreover, our estimates suggest the distributions are often fat-tailed, with critical implications for the probabilities of extreme SSEs. Evidence also suggests a potential role of policies in reducing the tail distributions.

### 2.1 Statistical model

Let us define the SSEs and their distribution. Following the notations of Lloyd-Smith et al. (2005), let *z*_*it*_ ∈ {0, 1, 2, …} denote the number of secondary cases an infected individual *i* has at time *t*. Then, given some threshold *<u>Z</u>*, an individual *i* is said to have caused SSE at time *t* if *z*_*it*_ ≥ *<u>Z</u>*. To make the estimation flexible, suppose the distribution for non-SSEs, *z*_*it*_ < *<u>Z</u>*, needs not follow the same distribution as those for SSEs.

In this paper, we consider a power law (or Pareto) distribution on the distribution of SSE. Denoting its exponent by *α*, the countercumulative distribution is

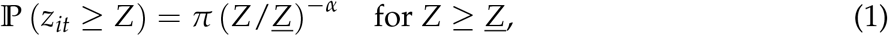

where *π* is the probability of SSEs. Notably, its mean and variance may not exist when *α* is sufficiently low: while its mean is 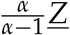 if *α* > 1, it is ∞ if *α* ≤ 1. While its variance is 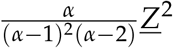 if *α* > 2, it is ∞ if *α* ≤ 2. In this paper, we formally call a distribution to be fat-tailed if *α* < 2 so that they have infinite variance. While non-existence of mean and variance may appear pathological, a number of socioeconomic and natural phenomenon such as city sizes (*α* ≈ 1), income (*α* ≈ 2), and earthquake energy (*α* ≈ 1) have tails well-approximated by this distribution as reviewed in the Introduction. A theoretical reason why this distribution could be relevant for airborne diseases is that the number of connections in social networks often follow a power law (Barabasi and Frangos, 2014).

This characteristics stands in contrast with the standard assumption in epidemiology literature that the full distribution of *z*_*it*_ follows a negative binomial (or Pascal) distribution^4^ with finite mean and variance. The negative binomial distribution has been estimated to fit the data better than Poisson or geometric distribution for SARS (Lloyd-Smith et al., 2005), and given its theoretical bases from branching model (e.g. Gay et al., 2004), it has been a standard distributional assumption in the epidemiology literature (e.g. Nishiura et al., 2017).

### 2.2 Data

This paper uses five datasets of recent coronavirus outbreaks for examining the distribution of SSEs: COVID-19 data from (i) across the world, (ii) Japan, and (iii) India, and (iv) SARS data, (v) MERS data.

i. **COVID-19 data from around the world:** this dataset contains clusters of infections found by a systematic review of academic articles and media reports, conducted by the Centre of the Mathematical Modelling of Infectious Diseases COVID-19 Working Group (Leclerc et al., 2020). The data are restricted to first generation of cases, and do not include subsequent cases from the infections. The data are continuously updated, and in this draft, we have used the data downloaded on June 3rd. There were a total of 227 clusters recorded.
ii. **COVID-19 data from Japan:** this dataset contains a number of secondary cases of 110 COVID-19 patients across 11 clusters in Japan until February 26th, 2020, reported in Nishiura et al. (2020). This survey was commissioned by the Ministry of Health, Labor, and Welfare of Japan to identify high risk transmission cases.
iii. **COVID-19 data from India:** this dataset contains the state-level data collected by the Ministry of Health and Family Welfare, and individual data collected by covid19india.org.^5^ We use the data downloaded on May 31st.
iv. **SARS from around the world:** this dataset contains 15 incidents of SSEs from SARS in 2003 that occured in Hong Kong, Beijing, Singapore, and Toronto, as gathered by Lloyd- Smith et al. (2005)^6^ through a review of 6 papers. The rate of community transmission was not generally high so that, for example, the infections with unknown route were only about 10 percent in the case of Beijing. The data consist of SSEs, defined by epidemiologists (Shen et al., 2004) as the cases with more than 8 secondary cases. For Singapore and Beijing, the contact-tracing data is available from Hsu et al. (2003) and Shen et al. (2004), respectively. When compare the fit to the negative binomial distribution, we compare the fit of power law to that of negative binomial using these contact tracing data.
v. **MERS from around the world:** this dataset contains MERS clusters reported up to August 31, 2013. The cases are classified as clusters when thee are linked epidemiologically. The data come from three published studies were used in Kucharski and Althaus (2015). Total of 116 clusters are recorded.

We use multiple data sets in order to examine the robustness of findings.^7^ Having multiple data sets can address each other’s weaknesses in data. While data based on media reports is broad, they may be skewed to capture extreme events; in contrast, data based on contact tracing may be reliable, but are restricted to small population. By using both, we can complement each data’s weaknesses.

### Estimation

The datasets report cumulative number of secondary cases, either ∑_*i*_ *z*_*it*_ (when a particular event may have had multiple primary cases) or ∑_*t*_ *z*_*it*_ (when an individual infects many others through multiple events over time). Denoting these cumulative numbers by *Z*, we consider this distribution for some *Z* ≥ *<u>Z</u>*^∗^. As discussed in Appendix A.1, we can interpret the estimates of this tail distribution as approximately the per-period and individual tail distribution and therefore map directly to the parameter of the SIR model in the next section. The thresholds for inclusion, *<u>Z</u>*^∗^, will be chosen to match the threshold for SSEs when possible, but also adjust for the sample size. For COVID-19 in the world, we apply *<u>Z</u>* = 40 to focus on the tail of the SSE distribution. For SARS, we apply *<u>Z</u>* = 8 as formally defined (Shen et al., 2004). For other samples, we apply *<u>Z</u>* = 2 because the sample size is limited.

To assess whether the distribution of *Z* follows the power law, we adopt the regression-based approach that is transparent and commonly used. If *Z* follows power law distribution, then by (1), the log of *Z* and the log of its underlying rank have a linear relationship: log *rank*(*Z*) = −*α* log *Z* + log(*Nπ<u>Z</u>*^*α*^). This is because, when there are *N* individuals, the expected ranking of a realized value *Z* is 𝔼 *rank*(*Z*) ∼ ℙ (*z* ≥ *Z*)*N* for moderately large *N*. Thus, when *N* is large, we obtain a consistent estimate of *α* by the following regression:

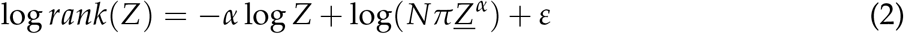

When *N* is not large, however, the estimate will exhibit a downward bias because log is a concave function and thus 𝔼 log *rank*(*Z*) < log 𝔼 *rank*(*Z*). While we present the analysis according to (2) in Figures 1 and 2 for expositional clarity, we also report the estimates with small sample bias correction proposed by Gabaix and Ibragimov (2011) in Appendix A.2.2.^8^ We also estimate using the maximum likelihood in Appendix A.2.2. Note that when there are ties (e.g. second and third largest had 10 infections), we assigned different values to each observation (e.g. assigning rank of 2 and 3 to each observation).

**Figure 2:**
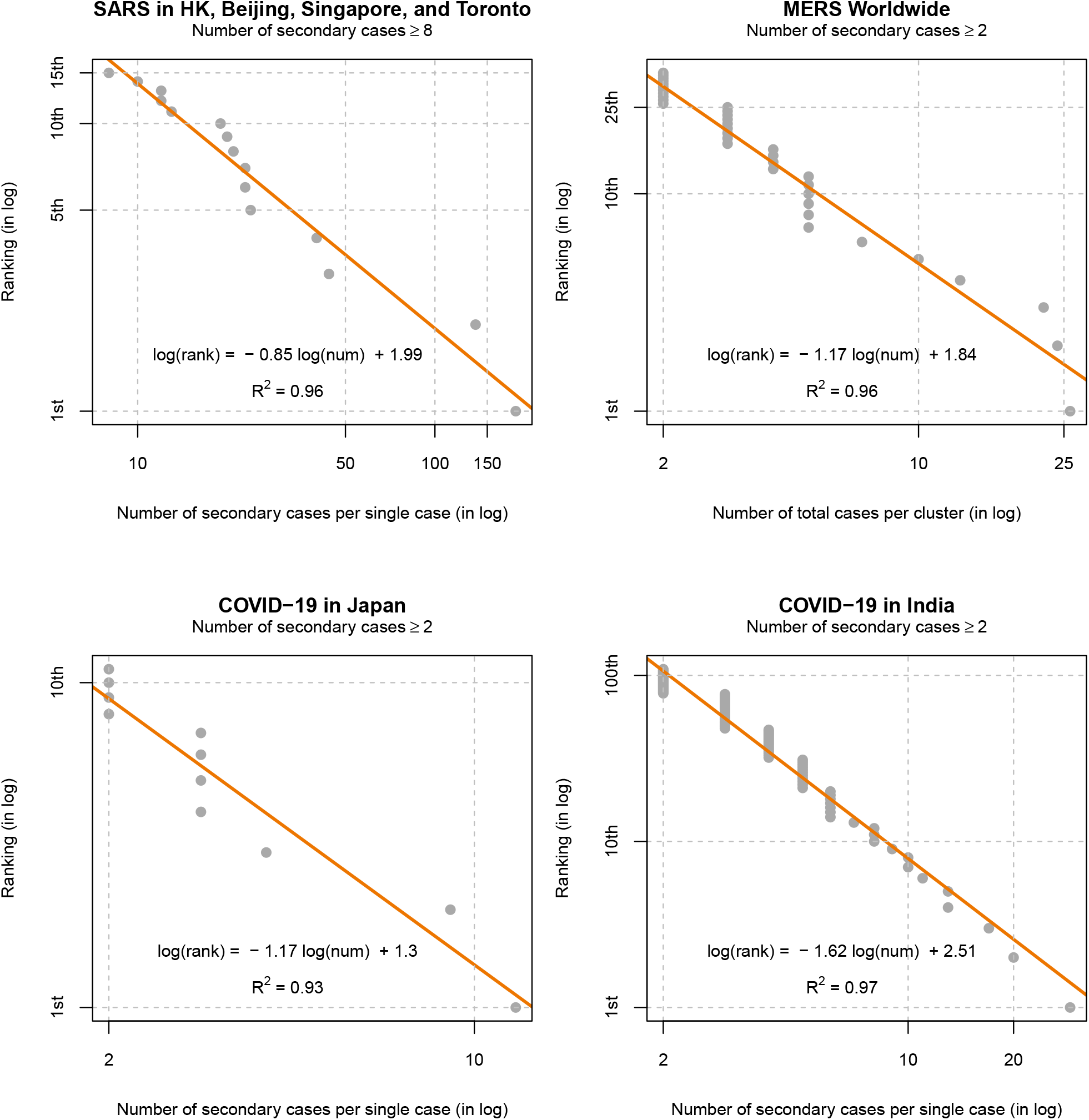
Log size vs log rank for COVID-19. *Notes*: Figure 2 plots the number of total cases per cluster (in log) and their ranks (in log) for MERS, and the number of total cases per cluster (in log) and their ranks (in log) for SARS and COVID-19 in Japan and India. The data for SARS are from Lloyd-Smith et al. (2005), and focus on SSEs defined to be the primary cases that have infected more than 8 secondary cases. The data for MERS come from Kucharski and Althaus (2015). The data for Japan comes from periods before February 26, 2020, reported in Nishiura et al. (2020). The data for India are until May 31, 2020, reported by the Ministry of Health and Family Welfare, and covid19india.org. The plots are restricted to be the cases larger than 2.

Next, we also compare the extent to which a power law distribution can approximate the distribution of SSEs adequately relative to the negative binomial distribution. First, we plot what the predicted log-log relationship in (2) would be given the estimated parameters of negative binomial distribution.^9^ Second, to quantify the predictive accuracy, we compute the ratio of likelihood of observing the actual data.

## 2.4 Results

Our analysis shows that the power law finely approximates the distribution of SSEs. Figure 1 visualizes this for COVID-19 from across the world, and Figure 2 for SARS, MERS, and COVID-19 in Japan and India. Their *R*^2^ range between 0.93 and 0.99, suggesting high levels of fit to the data. Because our focus is on upper-tail distribution, Figure 1 truncates below at the cluster size 40, Figure 2 truncates at 8 for SARS and at 2 for MERS and COVID-19 in India and Japan. Figure A.1 in Appendix presents a version of Figure 1 truncated below at 20.

In addition, the estimates of regression (2) suggest that the power law exponent, *α*, is below 2 and even close to 1. Table 1 summarizes the main findings. The estimated exponents near 1 suggest that extreme SSEs are not uncommon. For COVID-19 in Japan and India, the estimated exponents are larger than 1 but often below 2. Since applying the threshold of *<u>Z</u>*^∗^ = 2 is arguably too low, we must interpret out-of-sample extrapolation from these estimates with caution. When higher thresholds are applied, the estimated exponents tend to be higher. For example, when applying the threshold of *<u>Z</u>*^∗^ = 8 as in SARS 2003 to COVID-19 in India, the estimated exponent is 1.85 or 2.25. This pattern is already visible in Figure 2. Table A.1 in Appendix A.2.2 presents results using bias correction technique of Gabaix and Ibragimov (2011) as well as maximum likelihood. The results are very similar.

**Table 1:** Estimates of power law exponent 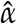 and their fit with data

|  | COVID-19 |  |  | SARS |  |  | MERS |
| --- | --- | --- | --- | --- | --- | --- | --- |
|  | World | Japan | India | World | Singapore | Beijing | World |
|  | (1) | (2) | (3) | (4) | (5) | (6) | (7) |
| $\hat{\alpha}$ | 1.07<br>(0.04) | 1.17<br>(0.10) | 1.62<br>(0.03) | 0.85<br>(0.06) | 0.75<br>(0.08) | 0.75<br>(0.06) | 1.17<br>(0.07) |
| $\underline{Z}$ | 40 | 2 | 2 | 8 | 2 | 2 | 2 |
| Obs. | 60 | 11 | 109 | 15 | 19 | 8 | 36 |
| $R^2$ | 0.98 | 0.93 | 0.97 | 0.96 | 0.91 | 0.94 | 0.96 |
| $\log_{10} LR$ | - | 11.39 | - | - | 19.51 | 8.04 | 40.89 |

*Notes:* Table 1 summarizes the estimates of power law exponent ( $\hat{\alpha}$ ) given as the coefficient of regression of log of number of infections (or size of clusters) on the log of their rankings. Heteroskedasticity-robust standard errors are reported in the parenthesis. $\underline{Z}$ denote the threshold number of infection to be included. $\log_{10}(LR)$ denotes “likelihood ratios”, expressed in the log with base 10, of probability of observing this realized data with power law distributions relative to that with estimated negative binomial distributions. Columns (1)-(3) report estimates for COVID-19; columns (4)-(6) for SARS, and column (7) for MERS.

Notably, the estimated exponent of India is higher than those of other data. There are two possible explanations. First, the lockdown policies in India have been implemented strictly relative to moderate approaches in Japan and some other parts of the world during the outbreaks. By discouraging and prohibiting large-scale gatherings, sometimes by police enforcement, they may have been successful at targeting SSEs. Second, contact tracing to ensure data reliability may have been more difficult in India until end of May than in Japan until end of February.^10^ While missing values will not generate any biases if the attritions were proportional to the number of infections, large gatherings may have dropped more than in Japan where the SSEs were found through contact tracing. Nonetheless, these estimates suggest that various environments and policies could decrease the risks of the extreme SSEs. This observation motivates our policy simulations to target SSEs.

Next, we compare the assumption of power law distribution relative to that of a negative binomial distribution. Figure 3 shows that the negative binomial distributions would predict that the extreme SSEs will be fewer than the observed distribution: while it predicts the overall probability of SSEs accurately, they suggest that, when they occur, they will not be too extreme in magnitude. Table 1 reports the relative likelihood, in logs, of observing the data given the estimated parameters. It shows that, under the estimated power law distribution relative to the estimated negative binomial distribution, it is 10^8^ − 10^20^ times more likely to observe the SARS data (10^40^ times more for MERS, and 10^11^ times more COVID-19 data in Japan). Such large differences emerge because the negative binomial distribution, given its implicit assumption of finite variance, suggests that the extreme SSEs are also extremely rare when estimated with entire data sets^11^. If our objective is to predict the overall incidents of infections parsimoniously, then negative binomial distribution is well-validated and theoretically founded (Lloyd-Smith et al., 2005).^12^ However, if our goal is to estimate the risks of extreme SSEs accurately, then using only two parameters with finite variance to estimate together with the entire distribution may be infeasible.

**Figure 3:**
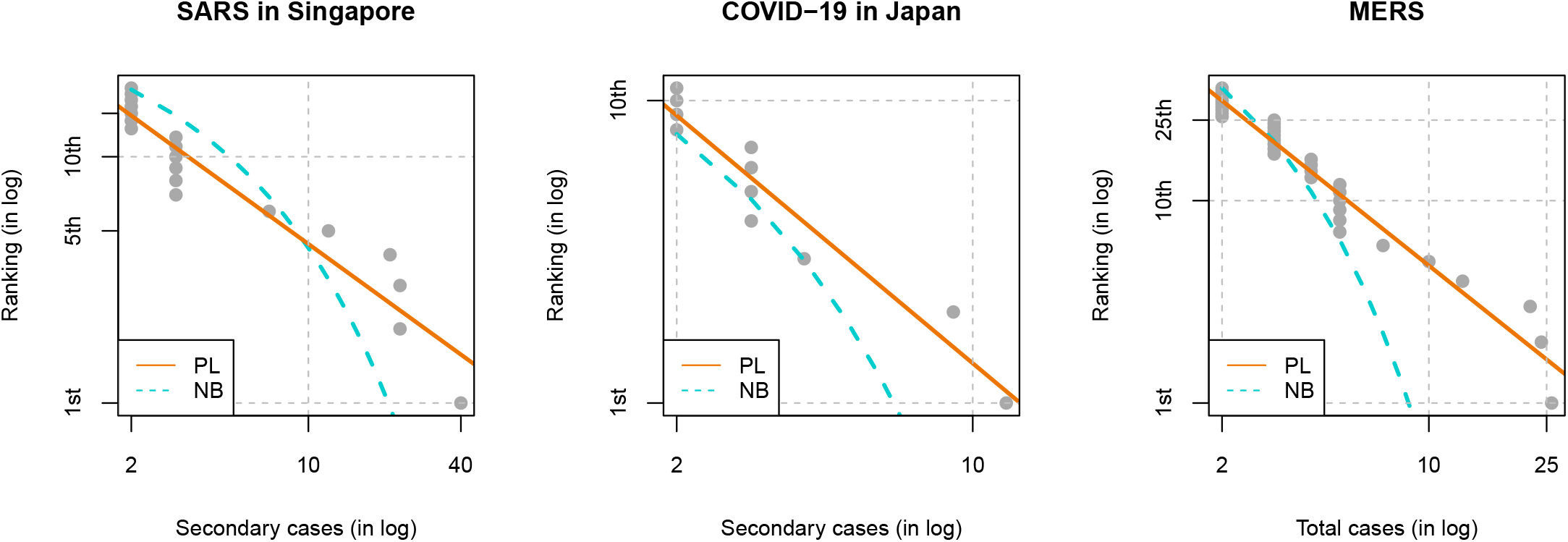
Comparison of power law and negative binomial distributions. *Notes*: Figure 3 plots the predicted ranking of infection cases given the estimated negative binomial (NB) distribution, in addition to the log-log plots and estimated power law (PL) distributions. The negative binomial distribution is parameterized by (*R, k*), where *R* is mean and *k* is the dispersion parameter with the variance being *R*(1 + *R*/*k*). The estimates for SARS Singapore come from our own estimates using the maximum likelihood (*R* = 0.88, *k* = 0.09); MERS come from the world (*R* = 0.47, *k* = 0.26) estimated in Kucharski and Althaus (2015); and COVID-19 in Japan were from our own estimates using the maximum likelihood (*R* = 0.56, *k* = 0.21). The estimates of Singapore is slightly different from Lloyd-Smith et al. (2005) because we pool all the samples.

These distributional assumptions have critical implications for the prediction of the extreme SSEs. Table 2 presents what magnitude top 1%, top 5%, and top 10% among SSEs will be given each estimates of the distribution. Given the estimates of the negative binomial distribution, even the top 1% of SSEs above 8 cases will be around the magnitude of 19-53. However, given a range of estimates from power law distribution, the top 1% could be as large as 569. Thus, it is no longer surprising that the largest reported case for COVID-19 will be over 1,000 people. In contrast, such incidents have vanishingly low chance under binomial distributions. Since the SSEs are rare, researchers will have to make inference about their distribution based some parametric methods. Scrutinizing such distributional assumptions along with the estimation of parameters themselves will be crucial in accurate prediction of risks of extreme SSEs.

**Table 2:** Probabilities of extreme SSEs under each distribution

|  | Power Law |  |  |  |  | Negative Binomial |  |  |
| --- | --- | --- | --- | --- | --- | --- | --- | --- |
| | $\alpha = 1.08$ | $\alpha = 1.1$ | $\alpha = 1.2$ | $\alpha = 1.5$ | $\alpha = 2$ | SARS | MERS | COVID-19 |
| 1% | 569 | 526 | 371 | 172 | 80 | 44 | 18 | 19 |
| 5% | 128 | 122 | 97 | 59 | 36 | 31 | 15 | 15 |
| 10% | 67 | 65 | 55 | 37 | 25 | 25 | 13 | 14 |

Notes: Table 2 shows the size of secondary cases at each quantile, top 1 percentile, 5 percentile, and 10 percentile, given each distributions. The negative binomial distribution’s estimates for SARS are from Singapore, for COVID-19 are from Japan, and for MARS is from around the world.

## 3 Theory

Motivated by the evidence, we extend an otherwise standard stochastic SIR model with a fat-tailed SSEs. Unlike with thin-tailed distributions, we show that idiosyncratic risks of SSEs induce aggregate uncertainties even when the infected population is large. We further show that the resulting uncertainties in infection rates have important implications for average epidemiological outcomes. Impacts of lockdown policies that target SSEs are discussed.

### 3.1 Stochastic SIR model with fat-tailed distribution

Suppose there are *i* = 1, …, *N* individuals, living in periods *t* = 1, 2, Infected individuals pass on and recover from infection in heterogeneous and uncertain ways. Let *β*_*it*_ denote the number of new infection in others an infected individual *i* makes at time *t*. Let *γ*_*it*_ ∈ {0, 1} denote the recovery/removal, where a person recovers (*γ*_*it*_ = 1) with probability *γ* ∈ [0, 1]. Note that, whereas *z*_*it*_ in Section 2 was a stochastic analogue of “effective” reproduction number, *β*_*it*_ here is such analogue of “basic reproduction number.” Assuming enough mixing in the population, these two models are related by 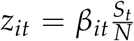, where *S*_*t*_ is a number of susceptible individuals in the population.

This model departs from other stochastic SIR models only mildly: we consider a fat-tailed, instead of thin-tailed, distribution of infection rates. Based onthe evidence, we consider a power law distribution of *β*_*it*_: its countercumulative distribution is given by

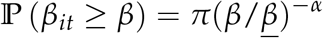

for the exponent *α* and a normalizing constant *<u>β</u>*, and *π* ∈ [0, 1] is the probability that *β* ≥ *<u>β</u>*. Note that the estimated exponent *α* can be mapped to this model, as discussed in Appendix A.1.

If we assume *β*_*it*_ is distributed according to exponential distribution or negative binomial distribution, we obtain a class of stochastic SIR models commonly studied in the epidemiological literature (see Britton (2010, 2018) for surveys). We will compare the evolution dynamics under this power law distribution against those under negative binomial distribution as commonly assumed, keeping the average basic reproduction number the same. To numerically implement this, we will introduce normalization to the distributions.

The evolution dynamics is described by the following system of stochastic difference equations. Writing the total number of infected and recovered/removed populations by *I*_*t*_ and *R*_*t*_, we have

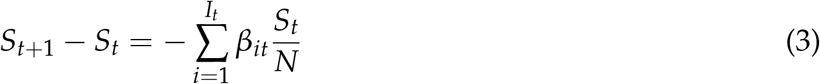

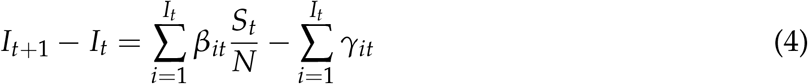

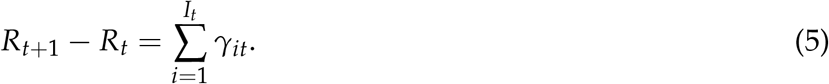

This system is a discrete-time and finite-population analogue of the continuous-time and continuous-population differential equation SIR models.

Parametrization: we parametrize the model as follows. The purpose of simulation is a proof of concept, rather than to provide a realistic numbers. We take the length of time to be one week. We set the sum of the recovery and the death rate per day is 1/18 following Wang et al. (2020), so that *γ* = 7/18. As a benchmark case, we set *α* = 1.1, which is the average from the our estimates from SARS and COVID-19 in Japan, but we explore several other parametrization, *α* ∈ {1.08, 1.2, 1.5, 2}. As documented in Nishiura et al. (2020), 75% of people did not infect others. We therefore set *π* = 0.25. This number is also in line with the evidence from SARS reported in Lloyd-Smith et al. (2005), in which 73% of cases were barely infectious. We choose <u>*β*</u>, which controls the mean of *β*_*it*_, so that the expected ℛ_0_ ≡ 𝔼*β*_*it*_/*γ* per day is 2.5, corresponding to the middle of the estimates obtained in Remuzzi and Remuzzi (2020). This leads us to choose <u>*β*</u> = 0.354 in the case of *α* = 1.1.

We will contrast the above model to a model in which *β*_*it*_ is distributed according to negative binomial, *β*_*it*_/*γ* ∼ negative binomial(ℛ_0_, *k*). The mean of this distribution is 𝔼*β*_*it*_/*γ* = ℛ_0_, ensuring that it has the same mean basic reproduction number as in the power law case, and the variance is ℛ_0_(1 + ℛ_0_/*k*). The smaller values of *k* indicate greater heterogeneity (larger variance). We use the estimates of SARS by Lloyd-Smith et al. (2005), *k* = 0.16. The mean is set to the same value as power law case, ℛ_0_ = 2.5,

### 3.2 Effects of fat-tailed distribution on uncertainty

Figure 4a shows 10 sample paths of infected population generated through the simulation of the model with *α* = 1.1. One can immediately see that even though all the simulation start from the same initial conditions under the same parameters, there is enormous uncertainty in the timing of the outbreak of the disease spread, the maximum number of infected, and the final number of susceptible population. The timing of outbreak is mainly determined by when SSEs occur. To illustrate the importance of a fat-tailed distribution, Figure 4b shows the same sample path but with a thin-tailed negative binomial distribution. In this case, as already 1,000 people are infected in the initial period, the CLT implies the aggregate variance is very small and the model is largely deterministic. This is consistent with Britton (2018). Britton (2018) shows that when the total population is as large as 1,000 or 10,000, the model quickly converges to the deterministic counterpart.

**Figure 4:**
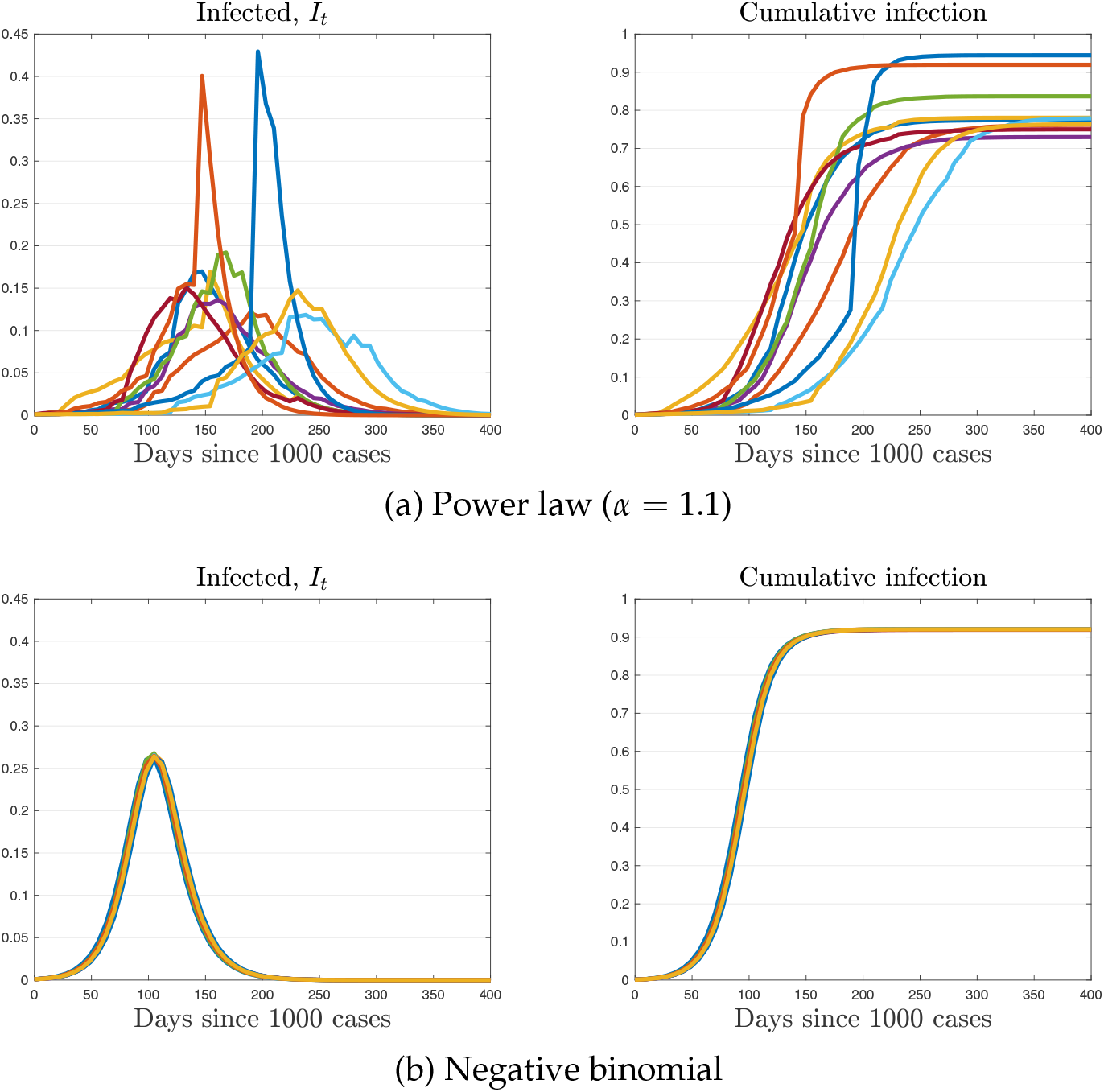
Ten sample paths from simulation. *Note:* Figure 4 plots 10 sample path of the number of infected population from simulation, in which we draw {*β*_*it*_, *γ*_*it*_} randomly every period in an i.i.d. manner. Figure 4a plots the case with power law distribution, and Figure 4b plots the case with negative binomial distribution.

Figure 5 compares the entire distribution of the number of cumulative infection (top-left), the herd immunity threshold (top-right), the peak number of infected (bottom-left), and the days it takes to infect 5% of population (bottom-right). The herd immunity threshold is defined as the cumulative number of infected at which the number of infected people is at its peak. The histogram contrast the case with power law distribution with *α* = 1.1 to the case with negative binomial distribution. It is again visible that uncertainty remains in all outcomes when the distribution of infection rate is fat-tailed. For example, the cumulative infection varies from 65% to 100% in the power law case, while the almost all simulation is concentrated around 92% in the case of negative binomial distribution.

**Figure 5:**
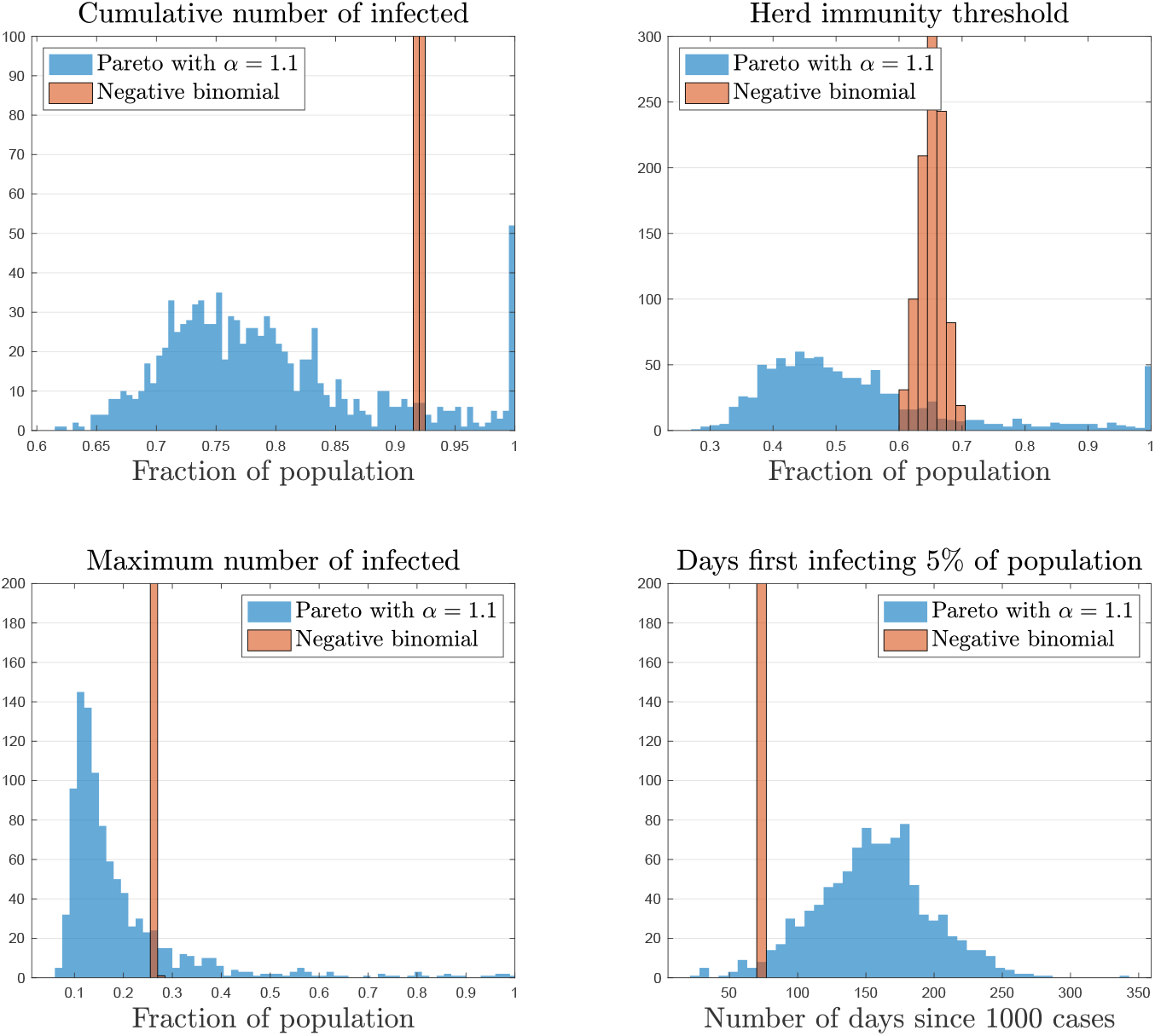
Histogram from 1000 simulation. *Note:* Figure 5 plots the histogram from 1000 simulations, in which we draw {*β*_*it*_, *γ*_*it*_} randomly every period in an i.i.d. manner. The cumulative number of infected is *S*_*T*_, where we take *T* = 204 weeks. The herd immunity threshold is given by the cumulative number of infected, at which the infection is at the peak. Formally, *S*_*t*_∗ where *t*^∗^ = arg max_*t*_ *I*_*t*_. The peak number of infected is max_*t*_ *I*_*t*_.

Table 4 further shows the summary statistics for the epidemiological outcomes for various power law tail parameters, *α*, as well as for negative binomial distribution. With fat-tails, i.e. *α* close to one, the range between 90th percentile and 10th percentile for all statistics is wide, but this range is substantially slower as the tail becomes thinner (*α* close to 2). For example, when *α* = 1.08 the peak infection rate can vary from 6% to 32% as we move from 10the percentile to 90th percentile. In contrast, when *α* = 2, the peak infection rate is concentrated at 26– 27%. Moreover, when *α* = 2, the model behaves similarly to the model with negative binomial distribution because the CLT applies to both cases.

**Table 3:**
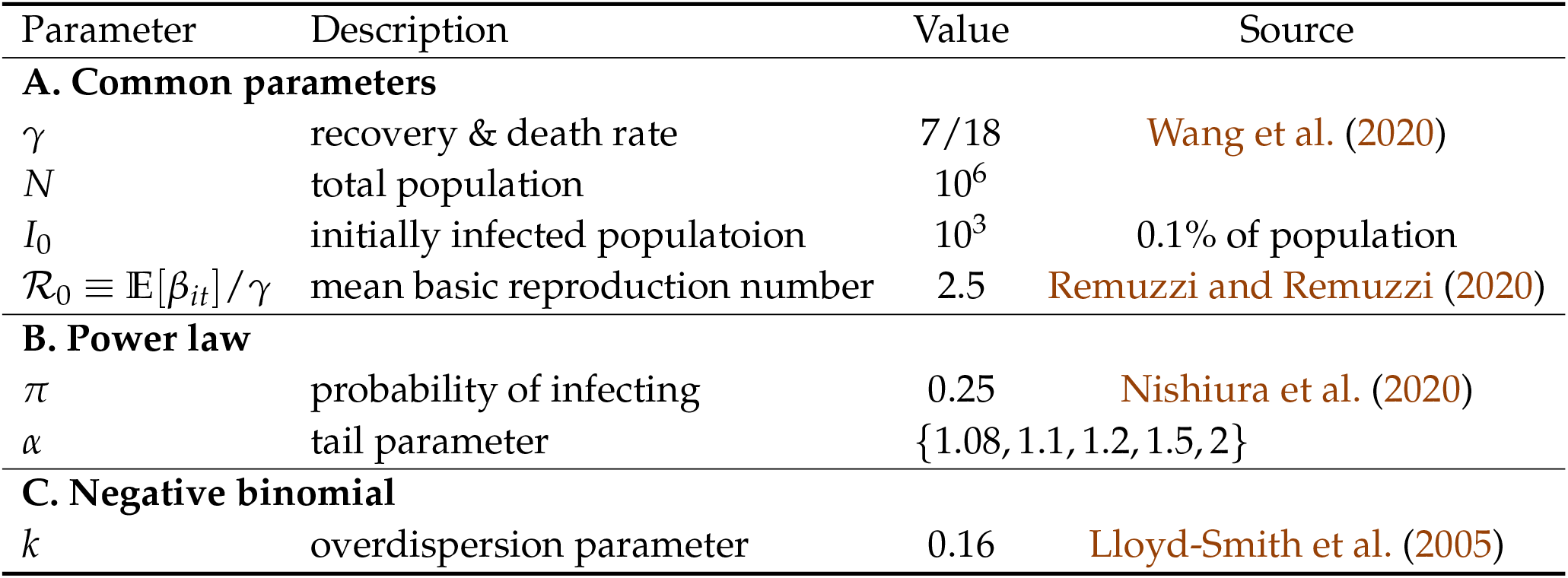
Parameter values

**Table 4:** Summary statistics for epidemiological outcomes

|  | Power law |  |  |  |  | Negative binomial |
| --- | --- | --- | --- | --- | --- | --- |
| | $\alpha = 1.08$ | $\alpha = 1.1$ | $\alpha = 1.2$ | $\alpha = 1.5$ | $\alpha = 2$ | |
| **1. Cumulative infected** |  |  |  |  |  |  |
| mean | 71% | 79% | 90% | 92% | 92% | 92% |
| 90th percentile | 88% | 93% | 94% | 92% | 92% | 92% |
| 50th percentile | 69% | 77% | 89% | 92% | 92% | 92% |
| 10th percentile | 58% | 70% | 87% | 91% | 92% | 92% |
| **2. Herd immunity threshold** |  |  |  |  |  |  |
| mean | 47% | 54% | 63% | 65% | 65% | 65% |
| 90th percentile | 70% | 80% | 72% | 70% | 68% | 68% |
| 50th percentile | 43% | 49% | 61% | 65% | 65% | 65% |
| 10th percentile | 30% | 38% | 55% | 60% | 62% | 63% |
| **3. Peak infection** |  |  |  |  |  |  |
| mean | 17% | 19% | 25% | 26% | 27% | 27% |
| 90th percentile | 32% | 34% | 31% | 27% | 27% | 27% |
| 50th percentile | 11% | 15% | 23% | 26% | 27% | 27% |
| 10th percentile | 6% | 10% | 20% | 25% | 26% | 26% |
| **4. Days infecting 5%** |  |  |  |  |  |  |
| mean (days) | 227 | 152 | 88 | 73 | 71 | 70 |
| 90th percentile | 343 | 203 | 105 | 77 | 77 | 70 |
| 50th percentile | 217 | 154 | 91 | 77 | 70 | 70 |
| 10th percentile | 119 | 98 | 70 | 70 | 70 | 70 |

*Note:* Table 4 shows the summary statistics from 1000 simulations for five different tail parameters for the case of power law distribution, and for the negative binomial distribution.

### 3.3 Effects of fat-tailed distribution on average

While our primary focus was the effect on the uncertainty of epidemiological outcomes, Figure 5 also shows significant effects on the mean. In particular, fat-tailed distribution also lowers cumulative infection, the herd immunity threshold, the peak infection, and delays the time it takes to infect 5% of population, *on average*. Why could such effects emerge?

To understand these effects, we consider a deterministic SIR model with continuous time and continuum of population. In such a textbook model, we consider the effect of small uncertainties (i.e. mean-preserving spread) in ℛ_0_. Such theoretical inquiry can shed light on the effect because the implication of fat-tailed distribution is essentially to introduce time-varying fluctuation in aggregate ℛ_0_. We can thus examine how the outcome changes by ℛ_0_, and invoke Jensen’s inequality to interpret the results.^13^

1. **Effect on cumulative infection:** note that the cumulatively infected population is given by 1 − *S*_∞_/*N*, where *S*_∞_ is the ultimate susceptible population as *t* → ∞. Taking the derivations shown in Harko et al. (2014), Moll (2020) or Toda (2020), *S*_∞_ satisfies the following equation:^14^

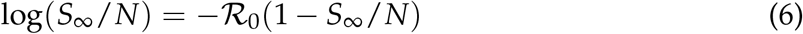 In Appendix B, we prove that *S*_∞_ is a convex function of ℛ_0_ if ℛ_0_ > 1.125,, which is likely to be met in SARS or COVID-19.^15^ Thus, the cumulative infection is concave in ℛ_0_, and the mean-preserving spread in ℛ_0_ lowers the cumulative infection.
2. **Effect on herd immunity threshold:** denoting the number of recovered/removed and infected population by *R*, the infection will stabilize when 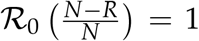. Rearranging this condition, the herd immunity threshold, *R*^∗^ is given by

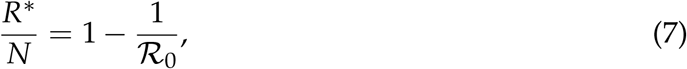

where ℛ_0_ ≡ *β*/*γ*. Thus, *R*^∗^ is concave in ℛ_0_. Thus, the mean-preserving spread in ℛ_0_ lowers the herd immunity threshold.
3. **Effect on timing of outbreak:** let us consider the time *t*^∗^ when some threshold of outbreak 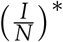 is reached. Supposing *S*/*N* ≈ 1 at the beginning of outbreak, *t*^∗^ satisfies

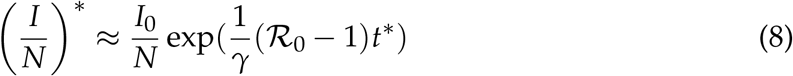 Thus, *t*^∗^ is convex in ℛ_0_, and the mean-preserving spread in ℛ_0_ delays the timing of the outbreak.
4. **Effect on peak infection rate:** the peak infection rate, denoted by 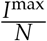, satisfies

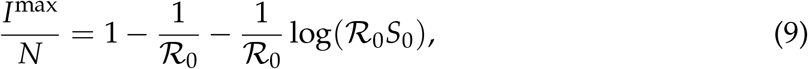

where *S*_0_ is initial susceptible population. We show in the Appendix that (9) implies that the peak infection, *I*^max^/*N*, is a concave function of ℛ_0_ if and only if 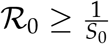 exp(0.5). If we let *S*_0_ ≈ 1, this implies ℛ_0_ ≥ exp(0.5) ≈ 1.65. This explains why we found a reduction in peak infection rate, as we have assumed ℛ_0_ = 2.5. Loosely speaking, since the peak infection rate is bounded above by one, it has to be concave for sufficiently high ℛ_0_.

Overall, we have found that the increase in the uncertainty over ℛ_0_ has effects similar to a decrease in the level of ℛ_0_. This is because the aggregate fluctuations in ℛ_0_ introduce negative correlation between the future infection and the future susceptible population. High value of today’s 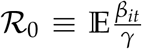 increases tomorrow’s infected population, *I*_*t*+1_, and decreases tomorrow’s susceptible population, *S*_*t*+1_. That is, *Cov*(*S*_*t*+1_, *I*_*t*+1_) < 0. Because the new infection tomorrow is a realization of *β*_*t*+1_ multiplied by the two (that is, 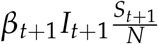) this negative correlation reduces the spread of the virus in the future on average, endogenously reducing the magnitude of the outbreak.

This interpretation also highlights the importance of intertemporal correlation of infection rates, *Cov*(*β*_*t*_, *β*_*t*+1_). When some individuals participate in events at infection-prone environments more frequently than others, the correlation will be positive. Such effects can lead to a sequence of clusters and an extremely rapid rise in infections (Cooper et al., 2019) that overwhelm the negative correlation between *S*_*t*+1_ and *I*_*t*+1_ highlighted above. On the other hand, when infections take place at residential environments (e.g. residential compound in Hong Kong for SARS, and dormitory in Singapore for COVID-19), then the infected person will be less likely to live in another residential location to spread the virus. In this case, the correlation will be negative. In this way, considering the correlation of infection rates across periods will be crucial.

Note that the mechanism we identified on herd immunity thresholds is distinct from the ones described in Gomes et al. (2020); Hébert-Dufresne et al. (2020); Britton et al. (2020). They note that when population has permanently heterogenous activity rate, which captures both the probability of infecting and being infected, the herd immunity can be achieved with lower threshold level of susceptible. They explain this because majority of “active” population becomes infected faster than the remaining population. Our mechanism does not hinge on the permanent heterogeneity in population, which could have been captured by *Cov*(*β*_*it*_, *β*_*it*+1_) = 1. The fat-tailed distribution in infection rate alone creates reduction in the required herd immunity rate in expectation.

### 3.4 Lockdown policy targeted at SSEs

How could the policymaker design the mitigation policies effectively if the distribution of infection rates is fat-tailed? Here, we concentrate our analysis on lockdown policy. Unlike the traditionally analyzed lockdown policy, we consider a policy that particularly targets SSEs. Specifically we assume that the policy can impose an upper bound on 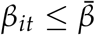 with probability *ϕ*. The probability *ϕ* is meant to capture some imperfection in enforcements or impossibility in closing some essential facilities such as hospitals and daycare. For tractability, we assume that the government locks down if the fraction of infected exceeds 5% of the population and maintain lockdown for 2 months. Nonetheless, our results are not sensitive to a particular parameter chosen. We also set 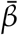 is 5 percentile of the infection rate.

While Table B.3 in Appendix presents results in detail, we briefly summarize the main results here. First, the lockdown policy reduces the mean of the peak infection rate, and the policy is more effective when the distribution features fatter tails. Second, the targeted lockdown policy is effective in reducing the volatility of the peak infection rate in the case that such risks exist in the first place. For example, consider the case with *α* = 1.1. Moving from no policy (*ϕ* = 0) to sufficiently targeted lockdown policy (*ϕ* = 0.8) reduces the 90th percentile of peak infection by 22%. In contrast, when *α* = 2 or with negative binomial distribution, the policy only reduces by 5% and 10%, respectively. Therefore the policy is particularly effective in mitigating the upward risk of overwhelming the medical capacity. This highlights that while the fat-tailed distribution induces the aggregate risk in the epidemiological dynamics, the government can partly remedy this by appropriately targeting the lockdown policy.

We conclude this section by discussing several modeling assumptions. First, we have assumed that {*β*_*it*_} is independently and identically distributed across individuals and over time. This may not be empirically true. For example, a person who was infected in a big party is more likely to go to a party in the next period. This introduces ex ante heterogeneities as discussed in (Gomes et al., 2020; Hébert-Dufresne et al., 2020; Britton et al., 2020), generating positive correlation in {*β*_*it*_} along the social network. Or, a person who tends to be a superspreader may be more likely to be a superspreader in the next period. This induces a positive correlation in {*β*_*it*_} over time. If the resulting cascading effect were large, then the average effects on the epidemiological outcomes we have found may be overturned. Second, we have exogenously imposed power law distributions without exploring underlying data generation mechanisms behind them. The natural next step is to provide a model in which individual infection rate is endogenously Pareto-distributed. We believe SIR models with social networks along the line of Pastor-Satorras and Vespignani (2001), Moreno et al. (2002), Castellano and Pastor-Satorras (2010), May and Lloyd (2001), Zhang et al. (2013), Gutin et al. (2020), and Akbarpour et al. (2020) are promising avenue to generate endogenous power law in individual infection rates.

## 4 Estimation methods

We began with the evidence that SSEs follow a power law distribution with fat tails in many settings, and showed that such distributions substantively change the predictions of SIR models. In this Section, we discuss the implications of power law distributions for estimating the effective reproduction number.

### 4.1 Limitations of sample means

Estimation of average reproduction numbers (ℛ_*t*_) has been the chief focus of empirical epidemiology research (e.g. Becker and Britton, 1999). Our estimates across five different data sets suggest that the exponent satisfies *α* ∈ (1, 2) in many occasions: that is, the infection rates have a finite mean but an infinite variance. Since the mean exists, by the Law of Large Numbers, the sample mean estimates (see e.g. Nishiura, 2007) that have been used in the epidemiology research will be consistent (i.e. converge to the true mean asymptotically) and also unbiased (i.e. its expectation equals the true mean with finite samples.)

Due to the infinite variance property, however, the sample mean will converge very slowly to the true mean because the classical CLT requires finite variance. Formally, while the convergence occurs at a rate 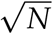 for distributions with finite variance, or thin tails, it occurs only at a rate 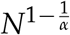 for the power law distributions with fat tails, *α* ∈ (1, 2) (Gabaix, 2011).^16^ Under distributions with infinite variance, or fat tails, the sample mean estimates could be far from the true mean with reasonable sample sizes, and their estimated 95 confidence intervals will be too tight. Figure 6 plots a Monte Carlo simulation of sample mean’s convergence property. For thin-tailed distributions such as the negative binomial distribution or the power law distribution with *α* = 2, even though the convergence is slow due to their very large variance, they still converge to the true mean reasonably under a few 1,000 observations. In contrast, with fat-tailed distributions such as power law distribution with *α* = 1.1 or *α* = 1.2, the sample mean will remain far from the true mean. Their sample mean estimates behave very differently as the sample size increases. Every so often, some extraordinarily high values occur that significantly raises the sample mean and its standard errors. When such extreme values are not occurring, the sample means gradually decrease. With thin tails, such extreme values are rare enough not to cause such sudden increase in sample means; however, with fat tails, the extreme values are not so rare.

**Figure 6:**
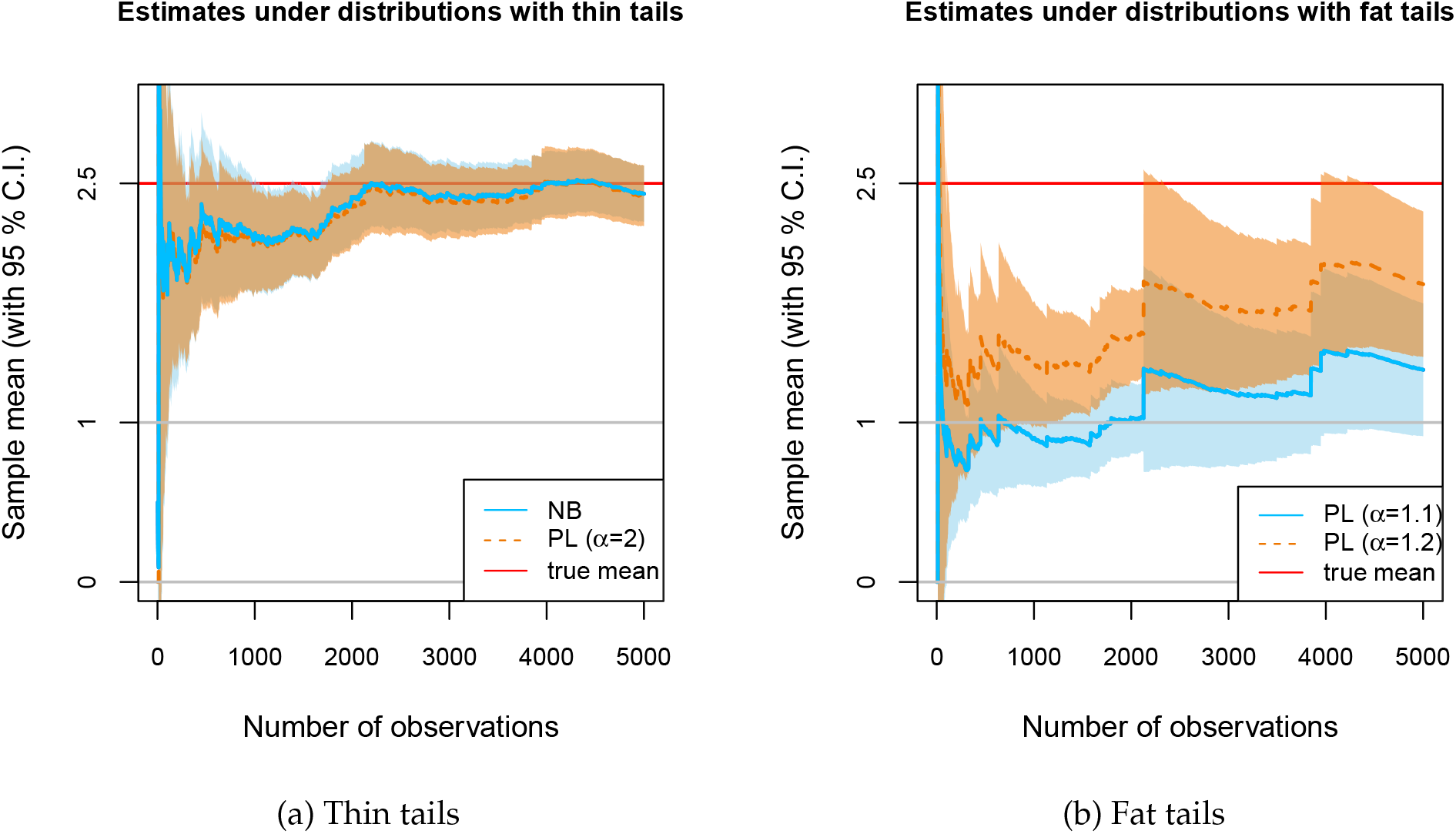
An example of sample mean estimates. *Notes*: Figure 6 depicts an example of sample mean estimates for thin-tailed and fat-tailed distributions. The draws of observations are simulated through the inverse-CDF method, where the identical uniform random variable is applied so that the sample means are comparable across four different distributions. All distributions are normalized to have the mean of 2.5. The negative binomial (NB) distribution has the dispersion parameter *k* = 0.16 taken from (Lloyd-Smith et al., 2005). The range of power law (PL) parameters is also taken from the empirical estimates.

### 4.2 Using power law exponents to improve inference

What methods could address the concerns that the sample mean may be empirically unstable? One approach may be to exclude some realizations as an outlier, and focus on subsamples without extreme values^17^. However, such analysis will neglect major source of risks even though extreme “outlier” SSEs may fit the power law distributions as shown in Figure 1. While estimating the mean of distributions with rare but extreme values has been notoriously difficult^18^, there are some approaches to address this formally.

With power law distributions, the estimates of exponent have information that can improve the estimation of the mean. Figure 7 shows that the exponents *α* can be estimated adequately with reasonable sample sizes.^19^ If *α* > 2, as may be the case for the India under strict lockdown, then one can have more confidence in the reliability of sample mean estimates. However, if *α* < 2, the sample mean may substantially differ from the true mean. At the least, one can be aware of the possibility.

**Figure 7:**
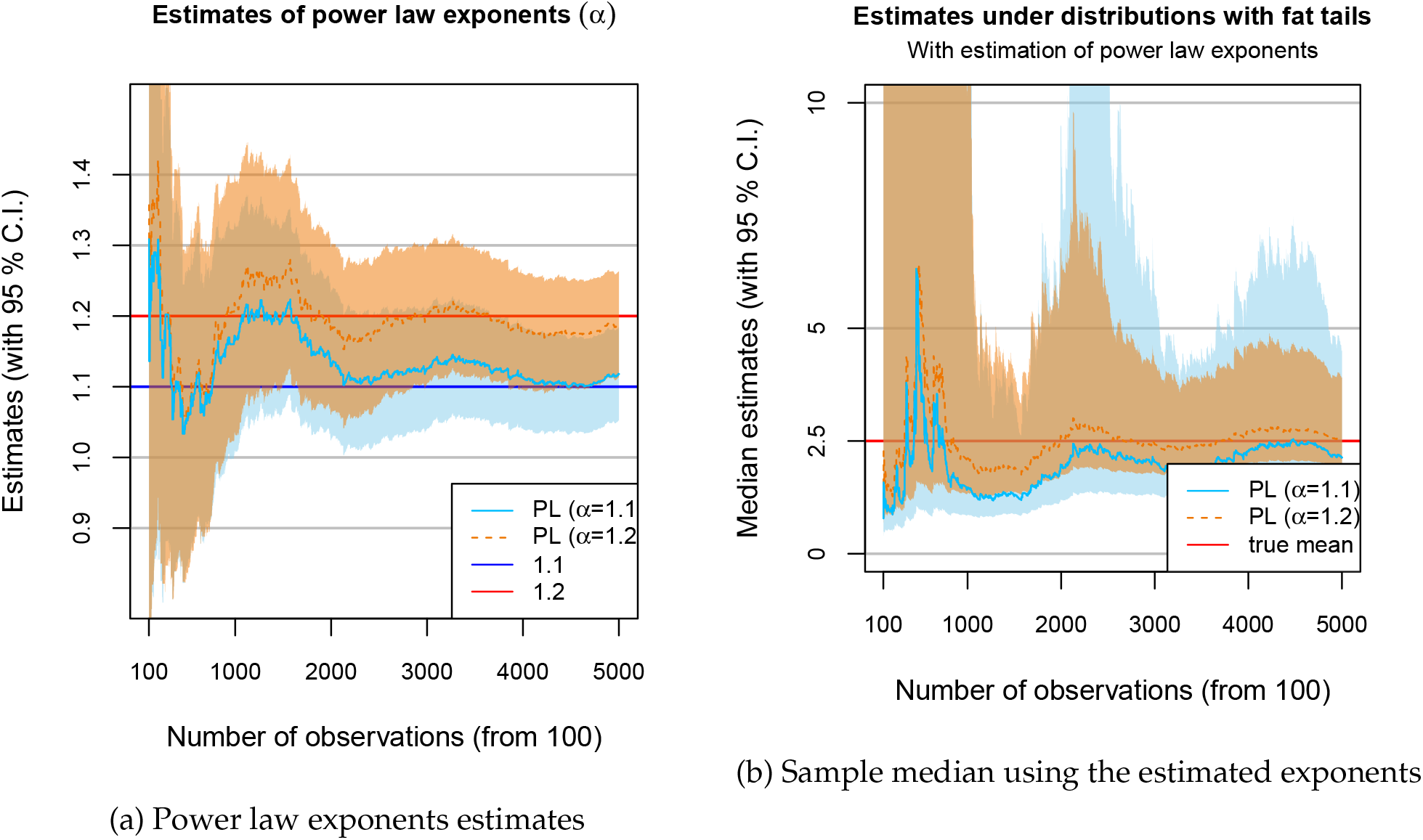
An example of “plug-in” estimates. *Notes*: Figure 7 plots the estimates of power law exponents and the resulting estimates of sample median, using the same data as in Figure 6. Note that while the number of observations contains all observations, the data points contributing to the estimates are only above some thresholds: only less than 25 percents of the data contribute to the estimation of the exponents.

One transparent approach is a “plug-in” method: to estimate the exponent 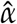, and plug into the formula of the mean 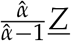. This method yields a valid 95 confidence intervals (C.I.) of the median^20^ since the estimated *α* has valid confidence intervals.^21^ Figure 7 shows the estimation results for the same data with *α* = 1.1, 1.2 as shown in Figure 6. First, while the sample mean in Figure 6 had substantially underestimated the mean, this estimated median is close to the true mean. Second, while the sample mean estimation imposed symmetry between lower and upper bounds of 95 percent confidence intervals, this estimate reflects the skewness of uncertainties: upward risks are much higher than downward risks because of the possibility of extreme events. Third, the standard errors are much larger, reflecting the inherent uncertainties given the limited sample sizes.^22^ Fourth, the estimates are more stable and robust to the extreme values^23^ than the sample mean estimates that have sudden jumps in the estimates after the extreme values.

Table 5 demonstrates the validity of the “plug-in” method through a simulation experiment. The table shows the comparison of the probability that the constructed 95% C.I. covers the true mean using the 1,000 Monte-Carlo simulation. When the estimate is unbiased and has correct standard errors, this coverage probability is 95%. When the power law exponent is close to one, the traditional “sample means” approach has the C.I. that covers the true mean only with 20-40% for all sample sizes. By contrast the “plug-in” method covers the true estimates close to 95%. As the tail becomes thinner toward *α* = 2, the difference between the two tends to disappear, with “sample mean” approach performing better some times. When the underlying distribution has fat-tails, however, estimation using the plug-in method is preferred.

**Table 5:**
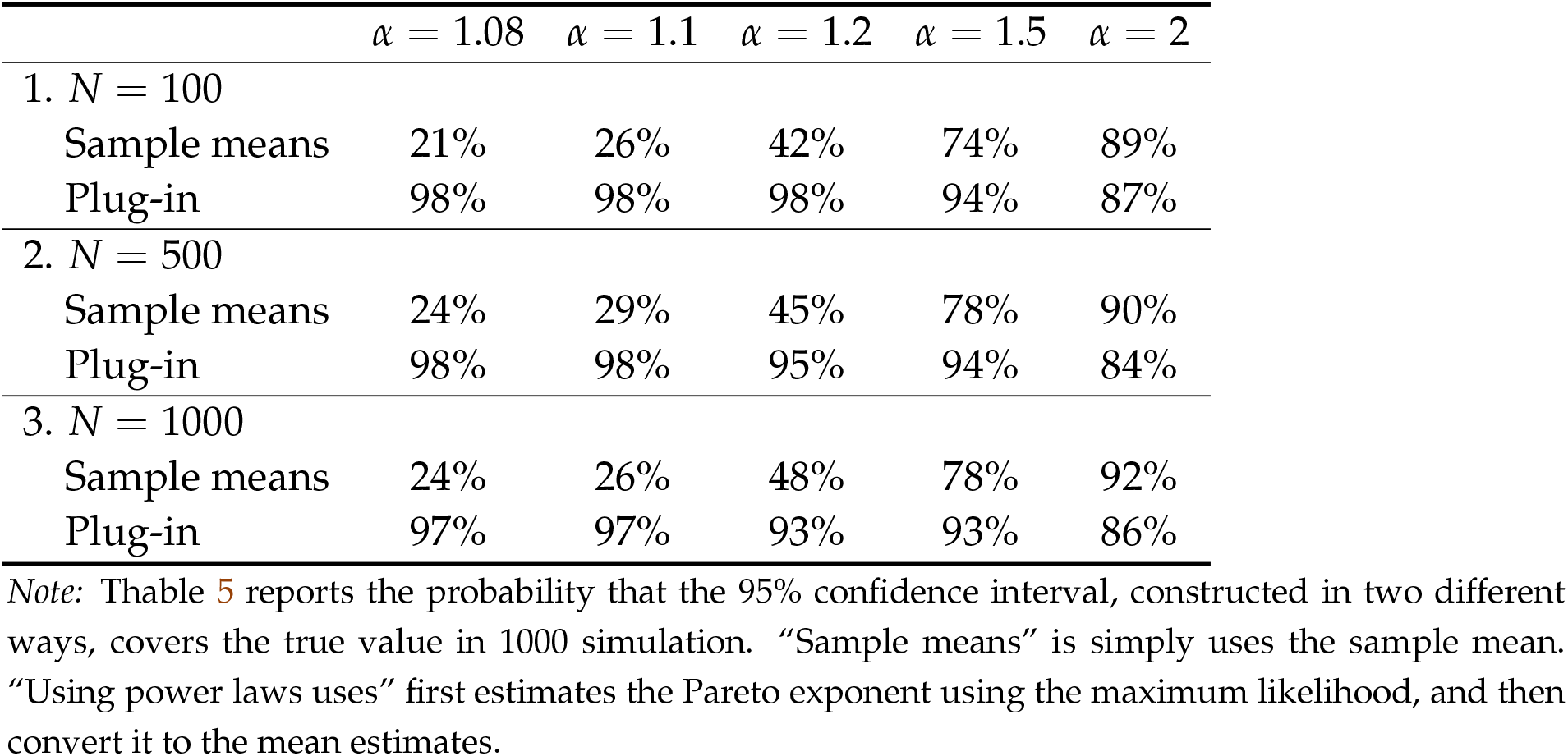
Coverage probability of 95% confidence interval

| | $\alpha = 1.08$ | $\alpha = 1.1$ | $\alpha = 1.2$ | $\alpha = 1.5$ | $\alpha = 2$ |
| --- | --- | --- | --- | --- | --- |
| 1. $N = 100$ | | | | | |
| Sample means | 21% | 26% | 42% | 74% | 89% |
| Plug-in | 98% | 98% | 98% | 94% | 87% |
| 2. $N = 500$ | | | | | |
| Sample means | 24% | 29% | 45% | 78% | 90% |
| Plug-in | 98% | 98% | 95% | 94% | 84% |
| 3. $N = 1000$ | | | | | |
| Sample means | 24% | 26% | 48% | 78% | 92% |
| Plug-in | 97% | 97% | 93% | 93% | 86% |

*Note:* Thable 5 reports the probability that the 95% confidence interval, constructed in two different ways, covers the true value in 1000 simulation. “Sample means” is simply uses the sample mean. “Using power laws uses” first estimates the Pareto exponent using the maximum likelihood, and then convert it to the mean estimates.

While the C.I. in the plug-in method has adequate coverage probabilities, it is often very large and possibly infinite. Figure 7 visualizes this. This large C.I. occurs especially when *α* ∼ 1 because the mean of a power law distribution is proportional to 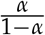 How could the policymakers plan their efforts do given such large uncertainty in ℛ_0_? Given the theoretical results in Section 3 that the epidemiological dynamics will be largely uncertain even when *α* ∼ 1 is perfectly known, we argue that applying the estimated ℛ_0_ into a deterministic SIR model will not lead to a reliable prediction. Instead of focusing on the mean, it will be more adequate and feasible to focus on the distribution of near-future infection outcomes. For example, using the estimated power law distribution, policymakers can compute the distribution of the future infection rate. The following analogy might be useful: in planning for natural disasters such as hurricanes and earthquakes, policymakers will not rely on the estimates of average rainfall or average seismic activity in the future; instead, they consider the probabilities of some extreme events, and propose plans contingent on realizations. Similar kinds of planning may be also constructive regarding preparation for future infection outbreaks.

To overcome data limitations, epidemiologists have developed a number of sophisticated methods such as backcalculation assuming Poisson distribution (Becker et al., 1991), and ways to account for imported cases. There are also a number of methods developed to account for fat-tailed distributions (see e.g. Stoyanov et al., 2010, for a survey), such as tail tempering (Kim et al., 2008) and separating the data into sub-groups (Toda and Walsh, 2015). In the future, it will be important to examine what power law distributions will imply about existing epidemiological methods, and how statistical techniques such as plug-in methods can be combined with epidemiological techniques to allow more reliable estimation of risks.

## 5 Conclusion: implications for COVID-19 pandemic

Most research on infection dynamics has focused on deterministic SIR models, and have estimated its key statistics, the *average* reproduction number (ℛ_0_). In contrast, some researchers have concentrated on SSEs, and estimated the *dispersion* of infection rates using negative binomial distributions. Nonetheless, stochastic SIR models based on estimated distributions have predicted that idiosyncratic uncertainties in SSEs would vanish when the infected population is large, and thus, the epidemiological dynamics will be largely predictable. In this paper, we have documented evidence from SARS, MERS, and COVID-19 that SSEs actually follow a power law distribution with the exponent *α* ∈ (1, 2): that is, their distributions have infinite variance rather than finite variance as with the negative binomial distributions. Our stochastic SIR model with these fat-tailed distributions have shown that idiosyncratic uncertainties in SSEs will persist even when the infected population is as large as 1,000, inducing major unpredictability in aggregate infection dynamics.

Since the currently infected population is estimated to be around 3 million in the COVID-19 pandemic,^24^ our analysis has immediate implications for policies of today. For statistical inference, the aggregate unpredictability suggests caution is warranted on drawing inferences about underlying epidemiological conditions from observed infection outcomes. First, large geographic variations in infections may be driven mostly by idiosyncratic factors, and not by fundamental socioeconomic factors. While many looked for underlying differences in public health practices to explain the variations, our model shows that these variations may be more adequately explained by the presence of a few, idiosyncratic SSEs. Second, existing stochastic models would suggest that, keeping the distribution of infection rates and pathological environments constant, recent infection trends can predict the future well. In contrast, our analysis shows that even when the average number of new infections may seem to have stabilized at a low level in recent weeks, subsequent waves can suddenly arrive in the future.

Such uncertainties in outbreak timing and magnitude introduce substantial socioeconomic difficulties, and measures to assess and mitigate such risks will be invaluable. Because the death rate is shown to increase when the medical capacity binds, the social cost of infection is a convex function of infection rates. In this sense, reducing uncertainties has social benefits. Furthermore, uncertainties can severely deter necessary investments and impede planning for reallocation and recovery from the pandemic shocks. To assess such risks, we can estimate the tail distributions to improve our inference on the average number. To address such risks, social distancing policies and individual efforts can focus on large physical gatherings in infection-prone environments. Our estimates suggest, like earthquakes, infection dynamics will be largely unpredictable. But unlike earthquakes, they are a consequence of social decisions, and efforts to reduce SSEs can significantly mitigate the uncertainty the society faces as a whole.

## Data Availability

All data is available upon request.

## Appendix

### A Empirical Appendix

#### A.1 Relating empirical distribution of *Z* to theoretical distribution of *β*_*it*_

In this paper, we have used the estimates from the data to simulate the evolution dynamics of the epidemiological model. The key step in our argument is that the tail distribution of ∑_*i*_ *z*_*it*_ or ∑_*t*_ *z*_*it*_, the *cumulative* “effective” number of infections, is equivalent to the tail distribution of *β*_*it*_, the *individual and per-period* “basic” number of infection. However, in general, this needs not hold: for example, even if *β*_*it*_ were normally distributed (i.e. thin tailed), *Z* may follow a *t*-distribution (i.e. fat-tailed). Under what conditions is our interpretation about the relationship between distribution of *Z* and distribution of *β*_*i*_ valid? Are they plausible in the settings of the coronaviruses?

To clarify this question, let us lay out a model. Formally, *Z* is a *mixture distribution* of the *weighted sum* of *β*_*it*_. Here, we provide notations for ∑_*t*_ *z*_*it*_ but the identical argument will also apply to ∑_*i*_ *z*_*it*_. Specifically, suppose *i* stays infected for 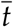 periods, and let the probability mass be 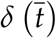. In the case of exponential decay as in the SIR model, 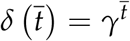. Denoting the counter-cumulative distribution of *Z*_*i*_ by ϕ, and that of *β*_*it*_ by *F*, we have

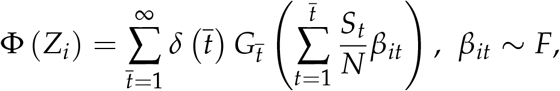

where 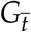 denotes the distribution 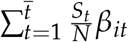.

##### A.1.1 Empirical evidence on causes of SSEs

First, we may be concerned that, even if ϕ is a power law distribution, *F* may not be a power law distribution. A counterexample is that a geometric Brownian motion with stochastic stopping time that follows exponential distribution can also generate power law distributions of the tail (Beare and Toda, 2020). That is, the tail property of ϕ needs not be due to tails of *F*: for ∑_*t*_ *z*_*it*_, it could also due to some individuals staying infectious for an extremely long periods. For ∑_*i*_ *z*_*it*_, it could also be due to some events having extremely high number of infected primary cases.

While we acknowledge such possibilities, we argue that for superspreaders or SSEs of the coronaviruses, the main mechanism of extremely high number of cumulative infection is primarily due to some extreme events at particular time *t*. Let us be concrete. If the counterexample’s reasoning were true for ∑_*t*_ *z*_*it*_, then a superspreader is someone who goes, for example, to a restaurant and infect two other people at time *t*, and then goes to a shopping mall and infects three other people at time *t* + 1, and then goes to meet her two friends and infect them, and so on. However, this interpretation is inconsistent with numerous anecdotes. Instead, a super-spreader infects many people because he attends a SSE that has infection-prone environment at a particular time *t*. Conferences, parties, religious gatherings, and sports gyms are a particular place that can infect many at the same time. Moreover, Nishiura et al. (2020) paper whose data we use has identified particular environment that has caused SSEs. This interpretation is important because, if the extremely high cumulative number of infection were due to some staying infectious for a long time or some events having extremely high number of primary cases, then our model’s prediction of sudden outbreak due to SSE is no longer a valid prediction.

##### A.1.2 Theoretical analysis on interpretation of exponents

Second, we may be concerned that the exponent of ϕ (*Z*_*i*_) may be different than the exponent of *F* (*β*_*iτ*_), even if both have tails that follow power laws. We use two steps to show that this is not a concern:

i. if a random variable has a power law distribution with exponent *α*, then its weighted sum also has a tail distribution that follows a power law with exponent *α* (see e.g. Jessen and Mikosch (2006) or Gabaix (2009)). Thus, neither summation over multiple periods nor the weights of 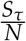 will change this.
ii. the tail property of distribution can be examined by considering 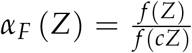 for some *c* /= 1 and taking its limit. In particular, if *F* has a power law distribution, then *α*_*F*_ (*Z*) = *c*^*α*^.^25^ Denoting the probability mass of 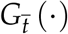 by 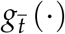, and the normalizing constant of each 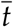 by 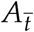,

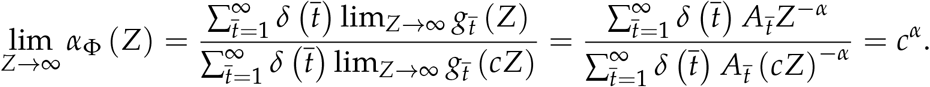

Thus, the exponent of ϕ (*Z*_*i*_) will be identical to the exponent of *F* (*β*_*iτ*_) asymptotically.

This discussion suggests that whenever possible, it is desirable to take the estimates from the tail end of the distribution instead of using moderate values of *Z*. For the COVID-19 from the world, the distributions are estimated from the very extreme tail. But when the sample size of SSEs is limited, choice of how many observations to include thus faces a bias-variance trade-off. Nonetheless, as many statistical theories are based on asymptotic results, these arguments show that it is theoretically founded to interpret the exponent of ϕ (*Z*_*i*_) as the exponent of *F* (*β*_*iτ*_), atleast given the data available.

#### A.2 Robustness

We present several robustness checks on our empirical results.

##### A.2.1 Figure 1 with a different cut-off

In Figure 1, we truncated the size of cluster from below at 40. Figure A.1 instead show results with a cut-off of 20. The fit is worse at the lower tail of the distribution, which suggests that the lower tail may not be approximated by power law distribution. This is a common feature among many examples. However, what matters for the existence of variance is the upper tail distribution, we do not think this is a concern. Moreover, given that the data partly come from media reports, the clusters of small sizes likely suffer from omission due to lack of media coverage.

##### A.2.2 Robustness of power law exponents estimates

Gabaix and Ibragimov (2011) show that an estimate of 2 is biased in a small sample and propose a simple bias correction method that replace the dependent variable with ln(*rank* − 1/2). Panel A of Table A.1 show the results with this bias correction method. The results are broadly very similar to our baseline results in Table 1.

Panel B of Table A.1 conduct another robustness check, where we estimate using the maximum likelihood. Again, the point estimates are overall similar to the baseline results, although standard errors are larger.

#### A.3 Additional Tables and Figures

Table A.2 shows several examples of superspreading events during COVID-19 pandemic.

**Figure A.1:**
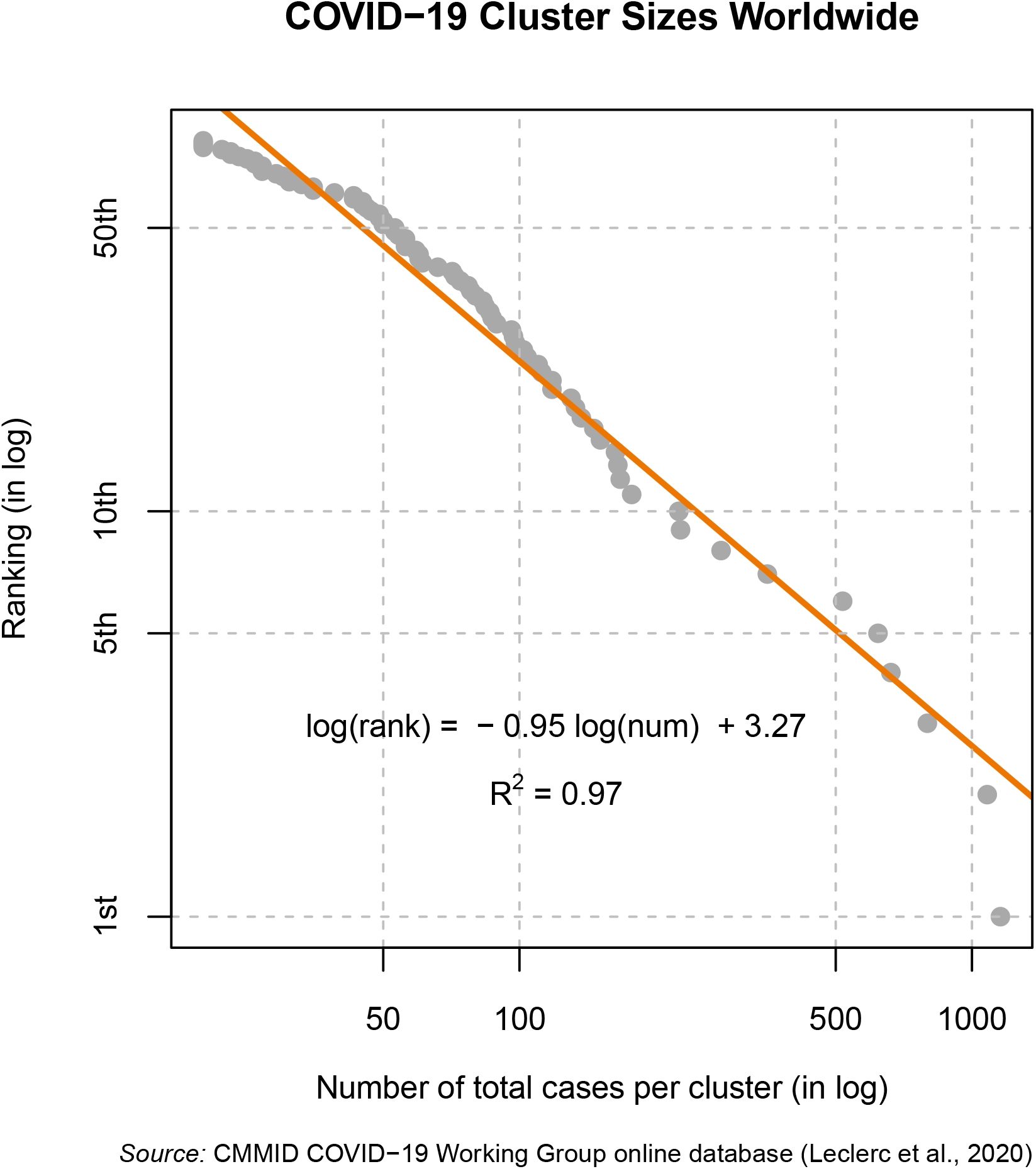
Log size vs log rank for Superspreading Events in SARS 2003. *Notes*: Figure A.1 plots the number of total cases per cluster (in log) and their ranks (in log) for COVID-19, last updated on June 3rd. It fits a linear regression for the clusters with size larger than 20. The data are collected by the Centre for the Mathematical Modelling of Infectious Diseases COVID-19 Working Group (Leclerc et al., 2020).

**Table A.1:**
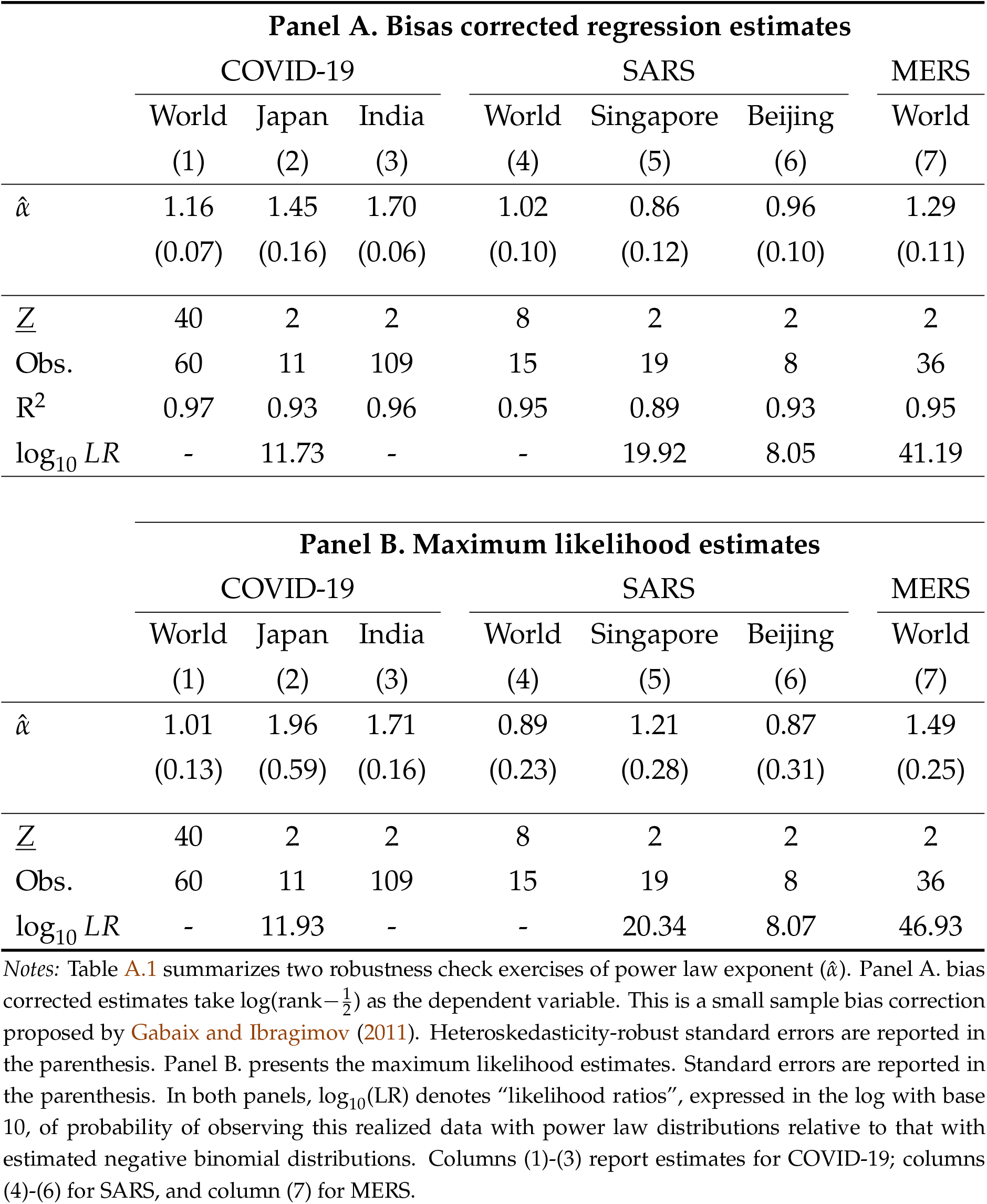
Estimates of power law exponent: robustness

**Table A.2:**
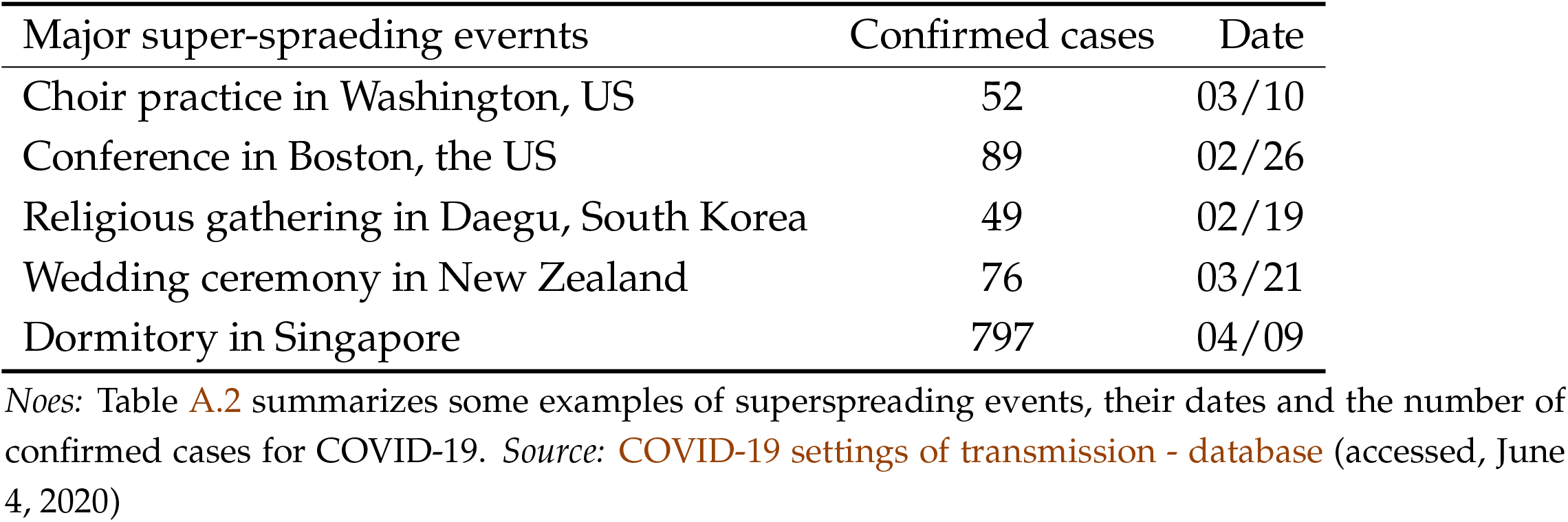
Examples of superspreading events

### B Theory Appendix

#### B.1 Proof that *S*_∞_ is convex in ℛ_0_ if 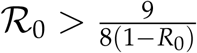

We show that *S*_∞_ is a concave function in ℛ_0_. Recall that *S*_∞_ is a solution to

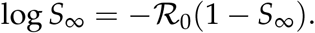

By the implicit function theorem,

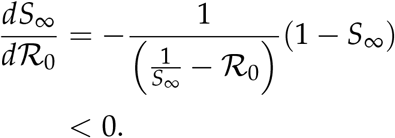

because *S*_∞_ < 1/ℛ_0_. Applying the implicit function theorem again,

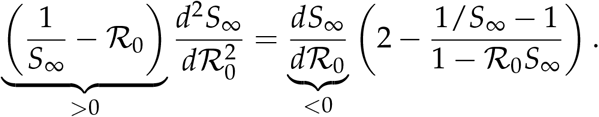

It remains to show that 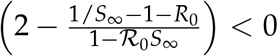 We can rewrite this as

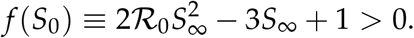

Note that *f* (·) is minimized at 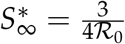. The minimum value is

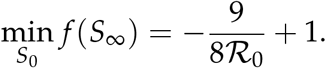

Therefore *f* (*S*_∞_) > 0 for all *S*_∞_ if and only if 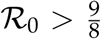. This implies that when 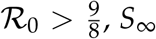, *S*_∞_ is a concave function of ℛ_0_.

#### B.2 Proof that *I*^max^ is concave in ℛ_0_ if and only if 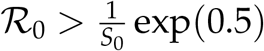

Recall that the peak infection rate is given by

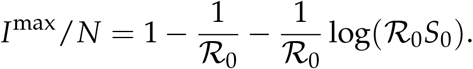

The derivative is

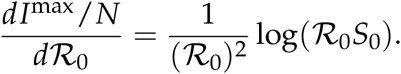

The second derivative is

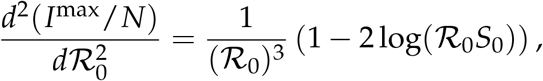

which is negative if and only if 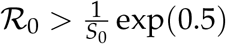.

#### B.3 Results for targeted lockdown policy experiment

Table B.3 shows the simulation results with lockdown policies targeted at SSEs. *ϕ* = 0 corresponds to no policy, *ϕ* = 0.4 (*ϕ* = 0.8) means that the government can prevent SSEs with 40% (80%) probability. As already discussed in the main text, when the distribution is fat-tailed, the targeted policy is not only effective in reducing the mean of the peak infection rate, but also its volatility (the interval between 90 percentile and 10 percentile).

**Table B.3:** Peak infection under targeted lockdown policy

|  | Power law |  |  |  |  | Negative |
| --- | --- | --- | --- | --- | --- | --- |
| | $\alpha = 1.08$ | $\alpha = 1.1$ | $\alpha = 1.2$ | $\alpha = 1.5$ | $\alpha = 2$ | binomial |
| **1. $\phi = 0$** |  |  |  |  |  |  |
| mean | 17% | 19% | 25% | 26% | 27% | 27% |
| 90th percentile | 32% | 34% | 31% | 27% | 27% | 27% |
| 50th percentile | 11% | 15% | 23% | 26% | 27% | 27% |
| 10th percentile | 6% | 10% | 20% | 25% | 26% | 26% |
| **2. $\phi = 0.4$** |  |  |  |  |  |  |
| mean | 11% | 12% | 16% | 21% | 24% | 21% |
| 90th percentile | 20% | 20% | 20% | 22% | 24% | 22% |
| 50th percentile | 7% | 8% | 14% | 21% | 24% | 21% |
| 10th percentile | 5% | 6% | 12% | 20% | 24% | 21% |
| **3. $\phi = 0.8$** |  |  |  |  |  |  |
| mean | 9% | 8% | 9% | 16% | 21% | 17% |
| 90th percentile | 14% | 12% | 11% | 17% | 22% | 17% |
| 50th percentile | 6% | 6% | 8% | 16% | 21% | 17% |
| 10th percentile | 5% | 5% | 7% | 15% | 21% | 16% |

*Note:* The table shows the summary statistics for peak infection rates from 1000 simulations with various policy parameters $\phi \in \{0, 0.4, 0.8\}$ , where $\phi$ denotes probability of limiting the size of SSEs to its five percentile.

## Footnotes

1 See Table A.2 in Appendix for a list of several examples.

2 In Appendix A.2.2, we also estimate the exponent with a small sample bias correction proposed by Gabaix and Ibragimov (2011), which shows the exponent is 1.16, and the *R*^2^ is 0.98. With maximum likelihood estimation, the exponent is 1.01.

3 For example, it is prohibitively costly to shut down daycare, but it is less costly to prevent a large concert.

4 Denoting its mean by *R* and dispersion parameter by *k*, the distribution is 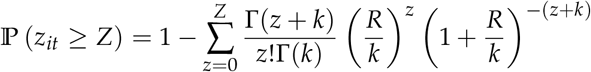 The variance of this distribution is 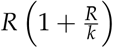. The distribution nests Poisson distribution (as *k* → ∞) and geometric distribution (when *k* = 1.)

5 https://www.kaggle.com/sudalairajkumar/covid19-in-india. covid19india.org is a volunteer-based organization that collects information from municipalities.

6 Even though Lloyd-Smith et al. (2005) had analyzed 6 other infectious diseases, SARS was the only one with sufficient sample sizes to permit reliable statistical analyses.

7 he infectious diseases considered here share some commonalities as SARS-CoV that causes SARS, MERS-CoV that causes MERS, and SARS-CoV-2 that causes COVID-19 are human coronaviruses transmitted through the air. They have some differences in terms of transmissibility, severity, fatality, and vulnerable groups (Petrosillo et al., 2020). But overall, as they are transmitted through the air, they are similar compared to other infectious diseases.

8 Their approach is to turn the dependent variable into 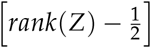 instead of log [*rank*(*Z*)]. We examine the performance of their bias correction method through a estimating regression given random variables generated from power law distributions. While their bias correction almost eliminates bias when *N* is moderately large, it has an upward bias of *α* whereas the equation (2) has a downward bias. The magnitude of bias is similar when *N* = 10 or *N* = 15. Thus, our preferred approach is to refer to both methods for robustness.

9 This approach stands in contrast with a common practice to plot the probability mass functions. Unlike such approaches where differences in tail densities are invisible since it is very close to zero, this approach highlights the differences in tail densities.

10 Concretely, there were only 248 cases of more than one secondary infections reported in the data among 27,890 primary cases in the data from India. That is, only 0.8 percents of primary cases were reported to have infected more than one persons. In contrast, there were 27 cases with more than one secondary infections among 110 primary cases in Japan. That is, 25 percent of primary cases were infectious. This difference in ration likely reflects the data collection quality than actual infection dynamics.

11 For example, the binomial distribution estimate suggests an incidence of 185 cases (residential infection in Hong Kong) only has a chance of 9.5 × 10^−10^ occurring for any single primary case.

12 Since the power law distribution is fitted only to SSEs, estimated power law distribution may fit the data better than the estimated negative binomial distribution that was meant to fit the entire data set. Rather than making such comparison, this estimation is intended to illustrate the magnitude of difference between the two distributional assumptions. Because of significant missing values for the low number of infections in the COVID-19 from across the world and India, we will not use the data sets for estimation of negative binomial distributions.

13 This assumes that ℛ_0_ is drawn at time 0, and stay constant thereafter for each simulation. This exercise is not exactly the same as our original SIR model because there ℛ_0_ fluctuates over time within a simulation. Thus this is for providing intuition, rather than a proof.

14 Here, we set the initially recovered population to zero, *R*_0_ = 0.

15 Numerically, we did not find any counterexample even when ℛ_0_ ∈ [1, 1.125].

16 For *α* = 1 exactly, the convergence will occur at rate ln *N*.

17 In Japan, the case of over 620 infections in the cruise ship Diamond Princess was excluded from all other analyses.

18 Consider, for example, a binary distribution of infection rates such that one infects *N* others with 1/*N* probability, and 0 others with 1 − 1/*N* probability. In this case, the true mean *R*_*t*_ = 1. Suppose a statistician observes 10 infected cases for each estimation. If *N* were 1,000, then with 99(≈ 0.999^10^) percent chance, nobody becomes infected so that 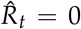, and the estimates’ confidence interval will be [0, 0]. But with less than 1 percent chance when any infection occurs, 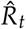 will be larger than 100. Thus, the 95 percent confidence interval contains the true mean in less than 1 percent of the time. To the best of our knowledge, there is no techniques that can help us completely avoid this problem given the fundamental constraint of small sample size.

19 The standard errors are computed by the maximum likelihood approach, as the linear regressions are known to underestimate the standard errors (see Gabaix and Ibragimov, 2011).

20 Note that the estimate corresponds to the median estimate because 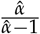 is a non-linear transformation of 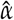.

21 To be more formal, the correct C.I. will be to consider the uncertainties with the mean of observations below *<u>Z</u>*. To focus on the uncertainty from upper tail, we construct the 95 percent C.I. from that of the estimate of $\alpha$ here.

22 When the number of observations is less than 1000, the estimated confidence interval of *α* contains values less than 1.0, turning the upper bound of the mean to be ∞. This does not mean that a correct expectation is ∞ infections in the near future, but that there is serious upward risks in infection rates.

23 This is because the estimation through log-likelihood will take the log of the realized value, instead of its level.

24 According to worldometers.info, the cumulative infection worldwide is 7 million, among which 4 million have already recovered or died, as of June 9, 2020.

25 This capture the essence of power laws – that whatever the value of *Z*, its frequency and frequency of *cZ* has the same ratio.

